# Timing of *M. tuberculosis* exposure explains variation in BCG effectiveness: A systematic review and meta-analysis

**DOI:** 10.1101/2020.11.26.20239533

**Authors:** James M Trauer, Andrew Kawai, Anna Coussens, Manjula Datta, Bridget M Williams, Emma S McBryde, Romain Ragonnet

## Abstract

**Background:** The variable efficacy observed in studies of BCG vaccination is incompletely explained by currently accepted hypotheses, such as latitudinal gradient in non-tuberculous mycobacteria exposure. We investigated heterogeneity in BCG vaccination in the context of participant demography, diagnostic approach and TB-related epidemiological context.

**Methods:** We updated previous systematic reviews of the effectiveness of BCG vaccination to 31^st^ December 2018. We employed an identical search strategy and inclusion/exclusion criteria to past reviews, but reclassified several studies and developed an alternative classification system.

**Results:** Of 21 included trials, those recruiting neonates and children aged under five were consistent in demonstrating considerable protection for several years. Trials in high-burden settings with shorter follow-up also showed considerable protection, as did most trials in settings of declining burden with longer follow-up. However, the few trials performed in high-burden settings with longer follow-up showed no protection, sometimes with higher case rates in the vaccinated than the controls in the later follow-up period.

**Conclusions:** The most plausible explanatory hypothesis is that BCG protects against TB that results from exposure shortly after vaccination. However, risk is equivalent or increased when exposure occurs later from vaccination, a phenomenon which is predominantly observed in adults in high-burden settings with longer follow-up. In settings of declining burden, most exposure occurs shortly following vaccination and the sustained protection thereafter represents continued protection against this early exposure. By contrast, in settings of continued intense transmission, initial protection subsequently declines due to repeated exposure to *M. tuberculosis* or other pathogens.

## Introduction

Tuberculosis (TB) is the world’s leading infectious disease killer (1), and Bacille Calmette-Guérin (BCG) is the only approved vaccine for its control. Global BCG coverage was 88% in 2019, close to the highest coverage of any vaccination (2). The substantial heterogeneity in the efficacy of BCG vaccination between studies has long been recognised, with study-level variables such as age, latitude and BCG strain able to explain some of this variation in meta-analyses unrestricted by age (3–5).

Past attempts to understand this heterogeneity have often started from the assumption that protection wanes with time from vaccination (6, 7). However, time since vaccination parallels immunological maturation and changing TB phenotype (8–10), which may lead to confounding. While retrospective national health data have shown that vaccine effectiveness can be sustained for >15 years (11, 12), multiple observational studies in low-burden settings have found that past history of BCG vaccination significantly increases risk in contacts of TB patients (13, 14).

These observations suggest that intensity or timing of *Mtb* exposure relative to age and time since vaccination may be important in determining vaccine effectiveness. However, no coherent theoretical framework has been proposed to explain the diversity of results and the adverse effects of BCG sometimes observed. Because of the markedly different disease phenotypes and onset timing with age of exposure (14, 15), we reviewed evidence for the efficacy of BCG vaccination with a focus on TB-related epidemiology, diagnostic approach, time from vaccination and age.

## Methods

### Reference management

We performed a systematic review of studies of the effect of BCG vaccination on TB disease, including sub-categories of TB disease, with search strategy and inclusion/exclusion criteria identical to Abubakar 2014 (5) and consistent with our PROSPERO-registered protocol (CRD42019119676), but with search dates extended to 31^st^ December 2018 (Section 2, Supplemental Appendix). Details of our search strategy are presented in Section 7 of the Supplemental Appendix. We focused on clinical trials as the highest form of evidence, but also reviewed cohort studies as a secondary level of evidence, as presented in the study profile (**Figure 1**). We considered studies comparing participants receiving their first BCG vaccination against unvaccinated controls, excluding trials of BCG revaccination, consistent with previous reviews. No further eligible trials were identified that had not been included in the earlier review, with the recent trial of placebo, H4:IC31 vaccine and BCG (which observed no TB cases, personal communication, Hatherill) excluded as a revaccination trial (16). All data extracted from studies published from 2009 to 2018 were reviewed by two authors (which pertains to cohort studies only).

**Figure 1.**
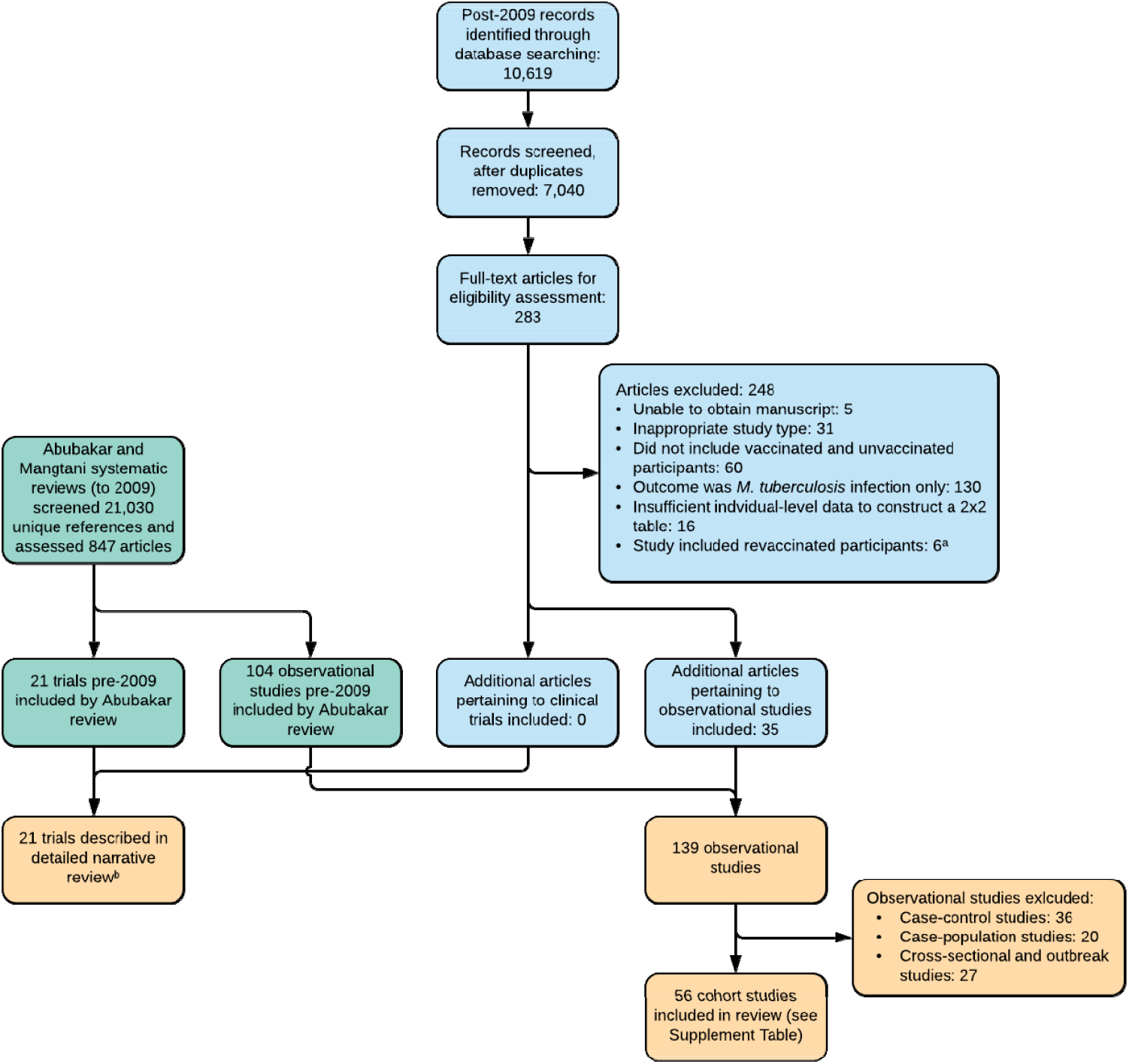
Modified PRISMA flow diagram. ^a^Includes two reports of one revaccination trial which observed zero cases of TB during follow-up. ^b^Studies in Chicago medical students, Chicago nursing students and New York infants not included in Mangtani review.

### Data extraction

We describe all trials in detail in our extended narrative review (Sections 3-11, Supplemental Appendix). Unlike previous reviews (4, 5, 17), we considered how age-specific reactivation profiles may be influenced by the background intensity of *Mtb* transmission and formulated a new classification for included trials and conceptual framework for integrating their results. Specifically, we considered participant age, background TB burden and duration of follow-up as the main factors in classifying included trials. Trials not exclusively recruiting neonates have previously been grouped according to study-level factors, including: stringency of latent TB infection (LTBI) testing, as either single or multiple tests; and age group, as either school-aged or other age. However, previous reviews classified several studies as “other age”, even though most participants were children, which we believe is misleading (4, 5). We also identified errors in the previous reviews (Section 2.3, Supplemental Appendix), including two major trials incorrectly assigned according to the authors’ classification system, with the differences between our results and those of the previous reviews confirmed by three authors blinded to each other’s assessment (JMT, AK, RR).

Because the Chengalpattu trial was the largest ever trial of BCG vaccination, was performed in a high-burden setting and was relatively recent, we also obtained unpublished estimates from this study disaggregated by age and time from vaccination. To illustrate the interacting effects of age and time from vaccination, we extracted data from reports of trials for which data could be disaggregated by both age at TB diagnosis and time from vaccination, in combination with the previously unpublished data from the Chengalpattu trial.

### Data synthesis and analysis

We used a hierarchical approach to estimating effect sizes, given the diverse approaches to presenting outcomes. Where possible we estimated effect sizes from information on person-years of follow-up and number of cases occurring in the vaccinated and unvaccinated groups. If this was unavailable, we estimated follow-up periods from the information provided (Section 2.2, Supplemental Appendix).

Meta-analysis was performed under our new classification with Stata version 16.1 (College Station, USA). The forest plot was generated using the random effects model of DerSimonian and Laird with the Mantel-Haenszel assessment of heterogeneity from the *metan* package, with pooled effect size estimates and confidence limits also presented using REML with Knapp-Hartung adjustment provided by Stata’s *meta* function.

Stata code for meta-analysis, and the data and Python 3.6 code for generating **Figure 2** and **Figure 3** are available at https://github.com/jtrauer/bcg_tb_context_review.

**Figure 2.**
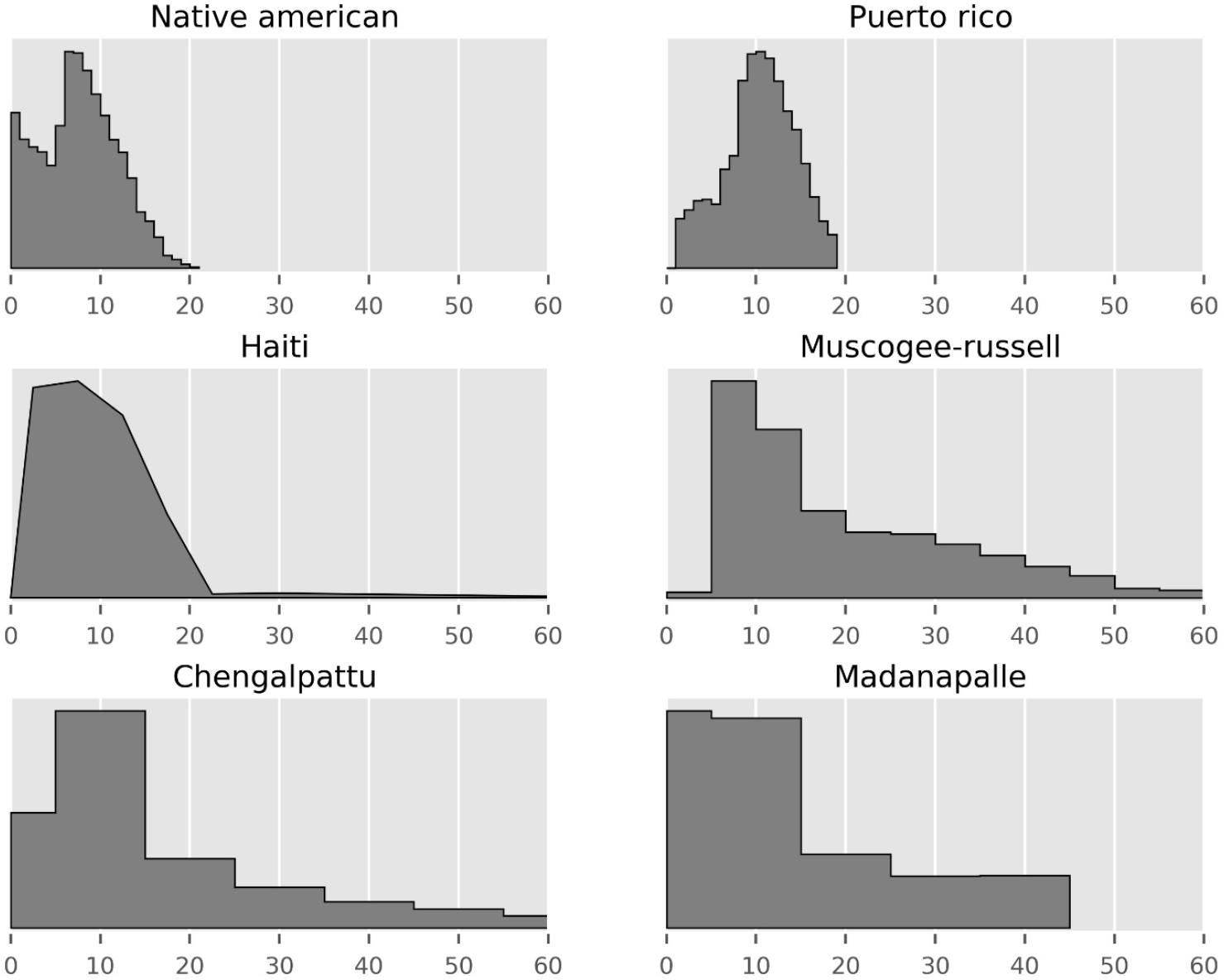
Age distribution (in years) of participants in studies for which these data were provided. Studies with very narrow age inclusion criteria not presented (i.e. five neonatal trials and English cities trial of children aged 14 to 15½ years). All age distributions normalised to the same maximum vertical height.

**Figure 3.**
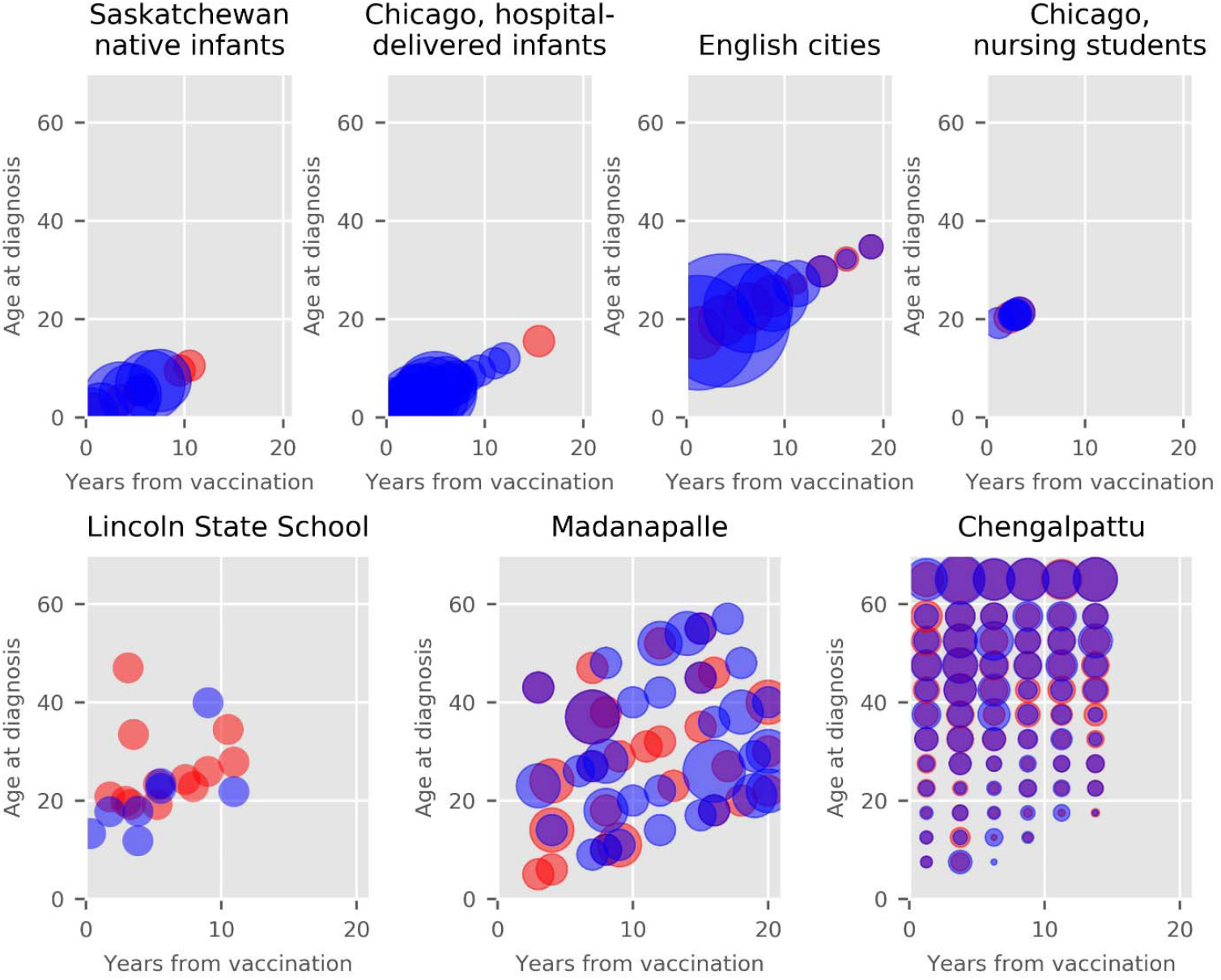
Distribution of cases of active TB occurring in the vaccinated (red) and unvaccinated (blue) populations in which timing of cases by age can be determined to within a five year interval. Size of circles proportional to the number of cases where multiple cases occurred within a time/age interval. First six studies assigned to vaccinated and control in a 1:1 ratio, while Chengalpattu assigned in ratio of 2 BCG: 1 control, with the size of the vaccinated circles halved to compensate for this effect. Madanapalle panel presents results for bacteriologically-confirmed cases only. First panel data obtained from Table IV of Ferguson 1949, second panel data obtained from Figure 1 of Rosenthal 1961, third panel data obtained from Table 3 of Medical Research Council 1972, fourth panel data obtained from Table 3 of Rosenthal 1963, fifth panel data obtained from Tables 1 and 2 of Bettag 1964, sixth panel data obtained from Table 5 of Frimodt-Möller 1973, seventh panel presents previously unpublished data.

## Results

Included studies are described in our detailed narrative review (Sections 3-11, Supplemental Appendix) and summarised as follows and in Table 2. Assessment of study quality according to standard criteria for the assessment of clinical trials has been undertaken in previous systematic reviews (Abubakar et al. 2013, Appendix 4) (5), and our assessments were in complete agreement with these findings.

**Table 1.** Proposed conceptual framework for understanding the effect of BCG vaccination. ^a^Early: approximately the first three to five years following vaccination; Late: greater than five years following vaccination.

| **Setting** | **Follow-up period<sup>a</sup>** | **Predominant reactivation profile** | **BCG effectiveness** |
| --- | --- | --- | --- |
| Declining burden | Early | Early progression from early infection/exposure | High |
| Declining burden | Late | Late progression from early infection/exposure | High |
| Sustained high burden | Early | Early progression from early infection/exposure | High |
| Sustained high burden | Late | Early progression from late infection/exposure | Low or adverse |

**Table 2.** Studies of BCG vaccination not restricted to neonates. ^a^TB cases were aged 15 to 44, such that “school” does not imply children. UK, United Kingdom; USA, United States of America. Chicago mental health patients trial (of participants aged up to 66 years) not included because only one unconfirmed case of TB was observed. ^b^Our assessment differs importantly from that of Mangtani et al. 2014. Italicised years indicate inferred/assumed dates/follow-up periods.

| Setting | Age profile | Recruitment start, recruitment end, follow-up end | Longest follow-up (years) | Epidemiological background | Latitude | TST number, dose | TST cut-off (mm) | Follow-up screening | Diagnostic confirmation | TB endpoint (authors' terminology) | Number recruited vaccinated, controls | TB cases in vaccinated, controls | Incidence rate ratio point estimate (confidence intervals) |
| --- | --- | --- | --- | --- | --- | --- | --- | --- | --- | --- | --- | --- | --- |
| Saskatchewan native infants, Canada | Neonates | 1933, 1945, 1947 | 14 | Rapidly declining following period of very high burden (53) | 50.8 N | None | N/A | X-ray | X-ray and clinical | TB | 306, 303 | 6, 29 | 0.19 (0.08 – 0.46) |
| Native infants, USA | Neonates | 1938, 1940, 1946 | 8 | Rapidly declining following period of very high burden (31) | 48.8 N and 43.2 N | None | N/A | X-ray, TST | Routine diagnosis after active follow-up ceased | Primary TB | 123, 139 | 4, 11 | 0.41 (0.13 – 1.29) |
| New York infants, USA | Neonates | 1933, 1944, 1944 | 11 | Rapidly declining following period of very high burden (54) | 40.7 N | None (for majority vaccinated before one month of age) | N/A | X-ray, TST | Microbiological or autopsy | None | 566, 528 | Not reported (TB-related deaths: 8, 8) | N/A (TB mortality: 0.93 (0.35 – 2.49)) |
| Chicago hospital-delivered infants, USA | Neonates | 1937, 1956, 1956 | 19 | Very high burden, then rapidly declining (55, 56) | 41.9 N | None | N/A | Clinical, TST, x-ray | Clinical, TST, x-ray, autopsy (x-ray sufficient for diagnosis) | TB | 5426, 4128 | 18, 63 | 0.22 (0.13 – 0.37) |
| Chicago household contact infants, USA | Neonates | 1940, 1955, 1955 | 15 | Very high burden, then rapidly declining (55, 56) | 41.9 N | None (if mother not the index) | N/A | Clinical, TST, x-ray | Clinical, TST, x-ray, autopsy (x-ray sufficient for diagnosis) | TB | 231, 220 | 3, 11 | 0.30 (0.08 – 1.07) |
| Mumbai infants, India | Neonates | 1972, 1972, 1975 | 2.5 | Very high burden (23) | 19.0 N | None | N/A | Clinical, TST | TST sufficient for diagnosis, although most TB diagnoses had x-ray changes | Primary TB | 396, 300 | 22, 27 | 0.63 (0.36 – 1.11) |
| Agra pre-school children, India | 0 to 5 years | 1979, 1979, 1984 | 5 | Very high burden (57) | 27.2 N | One-stage, 1 TU PPD-RT23 | <10 | X-ray, sputum examination | Not stated | Radiologically active or probably active TB | 1259, 1259 | 10, 25 | 0.40 (0.19 - 0.83) |
| Chicago housing project, USA | 0 to 12 years | 1942, 1955, 1955 | 13 | High burden (55, 56) | 41.8 N | Two-stage, Vollmer then 100 TU OT TST | Not stated | X-ray, TST | Unclear, presumed x-ray and clinical | Active pulmonary TB | 947, 944 | 0, 3 | 0.17 (0.01 - 3.32) |
| Jeremie population-wide, Haiti | All ages, see Figure 2 | 1965, 1966, 1969 | 3 | Very high burden (58) | 18.6 N | One-stage, 5 TU PPD-S | <6 | X-ray, TST | Microbiological | TB | 635, 338 | 1, 5 | 0.11 (0.01 - 0.91) |
| Puerto Rico children, USA | 1 to 18 years, see Figure 2 | 1949, 1951, 1969 | 19.8 | Declining but substantial following a period of very high burden (59) | 18.4 N | Predominantly (~76%) two-stage,<sup>b</sup> 1 then 10 TU PPD-RT19-20-21 | <6 | No active follow-up | Existing surveillance systems | TB | 50634, 27338 | 186, 141 | 0.71 (0.57 – 0.89) |
| Native Americans in four states, USA | 0 to 19 years, see Figure 1 | 1935, 1938, 1998 | 63 | Low and declining following a period of very high burden (60) | 32.2 N to 58.8 N | Two-stage, 1 then 250 TU PPD | Not stated | X-ray, TST | X-ray during first 12 years, then microbiological, clinical and other tests | Radiological evidence first 11 years, then by case definitions | 1551, 1457 | 84, 280 | 0.27 (0.21 - 0.34) |
| Georgia schools, USA | 6 to 17 years | 1947, 1947, 1967 | 20 | Low and declining following a period of high burden (61) | 32.5 N | Two-stage, 5 and 100 TU PPD-RT19-20-21 | <5 | TST | Clinical, microbiological, x-ray, TST | TB | 2498, 2341 | 5, 3 | 1.56 (0.37 – 6.54) |
| English cities, UK | 14 to 15½ years | 1950, 1952, 1972 | 20 | Low and declining following a period of high burden (62) | 51.5 N to 53.5 N | Two-stage, 3 and 100 TU OT | <5 | Clinical, TST, x-ray | Clinical, microbiological, pathological | TB | 13598, 12867 | 62, 248 | 0.23 (0.18 - 0.31) |
| Chicago mental health patients, USA | Adults up to 66 years | 1943, 1947, 1947 | 4 | High burden (63) | 41.9 N | Two-stage, 100 TU OT | Not stated | Not stated | Not stated | Pulmonary TB | 20, 15 | 0, 1 (unconfirmed case) | Not estimated |
| Chicago nursing students, USA | Presumed young adult | 1940, 1953, 1956 | 3 | High rates of exposure, higher in vaccinated (36) | 41.9 N | Two-stage, 2 and 10 TU OT | <6 | TST | Clinical, x-ray, other tests (x-ray sufficient for diagnosis) | TB | 231, 263 | 2, 5 | 0.45 (0.09 – 2.34) |
| Chicago medical students, USA | 20 to 37 years | 1939, 1952, 1964 | 4 | High rates of exposure (36) | 41.9 N | Two-stage, 2 and either 10 or 100 TU OT | <6 | TST | Clinical, x-ray, microbiological | TB | 324, 298 | 0, 3 | 0.15 (0.01 – 3.01) |
| Rand Mines, South Africa | 30.3 ± 10.3 years<sup>a</sup> | 1965, 1968, 1968 | 3.6 | Very high burden (64) | 26.2 S | None for most of trial, then one-stage for last eight months | Not stated | X-ray | X-ray | TB | 8317, 7997 | 29, 45 | 0.62 (0.39 – 0.99) |
| Muscogee and Russell Counties population-wide, USA | See Figure 2 | 1950, 1950, 1970 | 20 | Low and declining burden (61) | 32.5 N | One-stage, 5 TU PPD-RT-19-20-21 | <5 | None | Existing surveillance systems | TB | 16913, 17854 | 32, 36 | 0.94 (0.58 – 1.51) |
| Madanapalle population-wide, India | See Figure 2 | 1950, 1955, 1971 | 21 | Very high burden, rapidly declining due to active case finding (65) | 13.6 N | Predominantly one-stage (~95%), 5 TU PPD-RT-19-20-21<sup>b</sup> | <5 | X-ray | Clinical, x-ray, microbiological, other tests | Bacteriologically-confirmed TB | 5069, 5808 | 33, 47 | 0.81 (0.52 – 1.27) |
| Lincoln State School, USA | Not stated<sup>a</sup> | 1947, 1947, 1960 | 12 | Declining but substantial following a period of very high burden (66) | 40.2 N | Two-stage, 10 and 100 TU OT | Not stated | TST, x-ray | Clinical, microbiological, other tests | TB | 531, 494 | 12, 8 | 1.38 (0.56 – 3.38) |
| Chengalpattu population-wide, India | See Figure 2 | 1968, 1971, 1987 | 15 | Very high burden (67) | 12.7 N | One-stage, 3 TU PPD-S | <8 | X-ray, sputum examination | Culture-positive on ≥1 sputum | Culture-confirmed TB | 78693, 39025 | 380, 180 | 1.05 (0.88 – 1.25) |

### Neonatal vaccination trials

Of 21 included trials we identified, six recruited neonates only (18–23). A small minority of the participants in the Saskatchewan native infants and New York infants trials received oral vaccination, but most infants in these trials received parenteral vaccination. The trial in New York infants reported on TB-related mortality but not TB incidence, finding eight deaths in each of the vaccinated and control groups during its alternate assignment period (24). Although the trial undertaken in the lowest resource and likely highest transmission setting reported the lowest efficacy, estimates of protection were homogeneous, with most trials consistent with the pooled estimate of approximately 60% protection (4). Follow-up duration varied, although none followed participants into or beyond adolescence in a setting of continued intense transmission.

### Trials recruiting young children

Although the Agra trial was previously included with school-aged vaccination and is only reported very briefly, participants were aged <5 at entry (25). The point estimate of efficacy in this trial was 60% protection, consistent with estimates for neonates. The other trial recruiting the youngest children (previously classified as school-aged) observed three cases of TB, all among control participants, also consistent with high childhood efficacy (26).

### Reclassification of trials not restricted to young children

The categorisation of these studies as “school” or “other” age fails to convey that many studies previously categorised as “other” age predominantly recruited children. This often occurred because of an expansive population age pyramid and/or inclusion criteria, typically including negative LTBI testing, the prevalence of which declines with age, particularly in high-burden settings. A prime example is the population-wide Haiti trial, in which TST-negativity in adults was so rare that the protocol was modified to exclude participants >20, resulting in a predominantly paediatric cohort (**Figure 2**) (27).

### Trials recruiting broadly across paediatric age ranges

Three trials recruited across most or all paediatric age ranges to approximately 20 years (27–29). The Haitian trial observed participants for three years, with one case occurring in the vaccinated and five in the controls, suggesting good short-term protection in a high transmission setting (27). The large Puerto Rico trial (previously misclassified as non-stringent TST) was undertaken in a setting of rapidly declining but substantial burden, and showed modest efficacy (29, 30). The trial in native Americans was undertaken in a setting in which rates of disease rapidly declined to very low levels throughout most of follow-up, with sustained efficacy for >60 years (19, 28, 31).

### Paediatric/adolescent trials

The trial in Georgia schools recruited ages 6 to 17, with the small number of cases that occurred all accruing after six years of follow-up (30, 32). The large MRC-funded trial was undertaken in school children in large English cities and found high efficacy in a setting of rapidly declining burden (**Figure 3**) (33, 34).

### Trials in young adults with shorter follow-up

Three trials from Chicago of participants at high risk because of occupational or residential exposure risk were included in this category, along with one trial of participants at high occupational risk from a high-burden setting. The trial of mental health patients observed only one case of “bilateral minimal arrested tuberculosis” in 35 participants, providing little information. Two reports from Chicago describe BCG vaccination trials in students of nursing and medicine followed for the duration of their studies. Both were small and excluded from previous reviews for methodological limitations, with only eleven cases occurring across both trials (4, 35, 36). The trial in South African mine workers was also excluded from past reviews for including some participants who were TST-positive at baseline (6), but found fewer cases in the vaccinated over 3.6 years of follow-up (37).

### Trials recruiting across all ages with extended follow-up

The trial in Muscogee and Russell Counties, USA, achieved broad community participation in those aged >5 in a setting of low and declining transmission, finding slightly fewer cases in the vaccinated (30, 32). The trial in Lincoln State School describes an adult cohort followed in a high transmission setting and suggested a trend towards more cases in the vaccinated, particularly after five years of follow-up (**Figure 3**) (38).

The Chengalpattu trial was the largest BCG vaccination trial ever undertaken and was among the best reported, although it employed one-stage TST screening (39). It enrolled participants aged one month and above, followed for 15 years in a very high transmission setting and found slightly higher TB rates in the BCG-vaccinated, although protection was suggested in children (40). The Madanapalle trial (41) followed participants for 21 years in a very high transmission setting and focused on the end-point of smear-positive TB, which likely explains the low number of paediatric cases (**Figure 3**) (9). Although previously classified as employing stringent TST testing, one-stage testing with a cut-off of <5mm was used in around 95% of participants. A critical point of distinction between these two trials is that Madanapalle, unlike Chengalpattu, was linked to a successful case finding campaign that led to dramatic falls in TB mortality during the study period (42).

### Meta-analysis

Heterogeneity was not observed when studies were grouped according to our classification (Figure 4). The pooled estimate for the incidence rate ratio of BCG vaccination in trials of neonates and young children was 0.26 (95%CI 0.17, 0.35). For other paediatric studies in high-burden settings, we refer readers to the descriptions of the original trials. For paediatric studies with longer follow-up in settings of declining burden the incidence rate ratio was 0.25 (95%CI 0.20, 0.30). Trials of adults in high-burden settings with short follow-up durations were methodologically heterogeneous, but the pooled estimate suggested benefit, with an incidence rate ratio of 0.59 (95%CI 0.31, 0.87). Trials of adults with longer follow-up duration were all consistent with a null overall effect, with marginal protection was suggested in settings of declining burden and the Chengalpattu trial dominating the effect estimate in high-burden settings.

**Figure 4.**
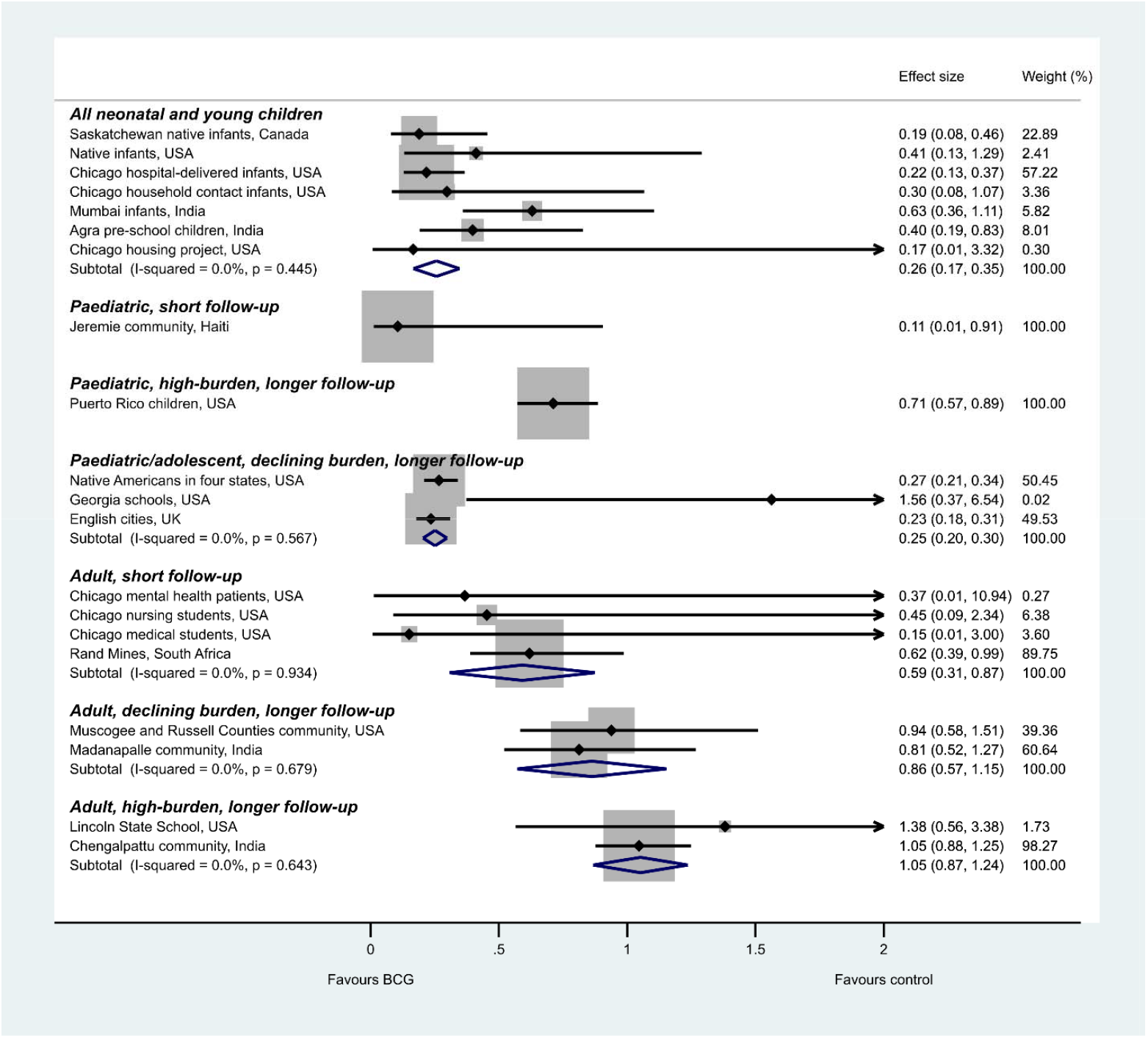
Forest plot of TB incidence rate ratios from trials of BCG vaccination by participant demographics and background epidemiology. Pooled effects are from random effects meta-analysis. Pooled estimates with confidence limits when using REML model with Stata 16.1 and Knapp-Hartung adjustments were: All neonatal and young children: 0.27 (0.15, 0.40); Paediatric/adolescent, declining burden, longer follow-up: 0.25 (0.17, 0.33); Adult, short follow-up: 0.59 (0.42, 0.77); Adult, declining burden, longer follow-up: 0.86 (0.08, 1.64), Adult, high-burden, longer follow-up: 1.05 (0.50, 1.61). New York infants trial not presented because only the outcome of TB-related deaths was reported from this trial.

### Cohort studies

Very few cohort studies were undertaken in high-burden settings with extended follow-up, and none disaggregated rates by both age and time (Table 1, Supplemental Appendix). Many cohort studies were undertaken in settings of declining burden with short follow-up durations, with most suggesting high efficacy. Several studies in settings of declining burden (France, Norway, Greenland) with extended periods of follow-up (>15 years) supported a sustained protective effect. However, several cohort studies with shorter follow-up duration in higher exposure settings suggested no effect (either high-burden countries, some studies of TB contacts or groups with occupational exposure risk).

## Discussion

Few trials of BCG vaccination followed participants for more than five years post-vaccination, but of those that did, settings with sustained exposure risk showed lower effectiveness than those in which risk predominantly accrued shortly after vaccination. Therefore, we propose that timing of *Mtb* exposure, rather than timing of TB disease, should be the main consideration in understanding the differences in BCG efficacy between settings (**Table 1**). In settings of rapidly declining TB burden, exposure is less likely further from vaccination and the effect of BCG in preparing the participant’s immune system for early exposure is robust. By contrast, in sustained high-burden settings, participants are more likely to be exposed after a longer period from vaccination when BCG efficacy has waned or after re-exposure has occurred. This interpretation implies that BCG may have little effect or could increase longer term-risk in settings of persistent exposure.

None of the eight studies of neonates or young children followed a substantial cohort of participants into the high-risk adolescent period in a high-transmission setting, but all were consistent with significant protection in early life. Among paediatric and adolescent trials, two larger studies in settings of declining burden showed substantial protection and one suggested short-term protection in a high-burden setting. In adults, four high-quality studies with long-term follow-up have been undertaken, with the two performed in high transmission settings suggesting an null or adverse effect (38, 39), while one in a low transmission setting (30, 32) and one linked to case finding (41) found modest protection. Methodologically heterogeneous trials with short follow-up of participants were also consistent with short-term protection in adults.

Included trials predominantly considered persons without prior *Mtb* sensitisation; however, the background intensity of *Mtb* transmission remains relevant to the relative importance of early and late reactivation (43). In settings of declining burden, TB episodes many years from vaccination are more likely to represent progression of infection acquired shortly following vaccination. Conversely, in high-burden settings, late-presenting episodes may result from later exposure. Under this conceptual framework, trials in both low- and high-burden settings will show high initial effectiveness, whereas later protection is dependent on transmission intensity. As late reactivation is commoner in adolescence and adulthood (14), these effects will be less apparent in studies that do not follow participants passing into adulthood. The recent BCG and H4:IC31 revaccination trial was conducted in adolescents in a high-burden setting. Both BCG and H4:IC31 boosting in those previously BCG vaccinated decreased persistent *Mtb* sensitisation, consistent with short-term protection from infection (16).

A non-significant effect in studies enrolling across a broad range of ages should not be interpreted as lesser protection in older participants, because such studies may include substantial cohorts of young children and follow-up invariably emphasises the early post-vaccination period. Therefore, it is more likely that protection is present in young children, but offset by adverse effects in adults in the late post-vaccination period. An example is the Chengalpattu trial which reported considerable protection in children aged <15 during the first 12.5 years of follow-up (40), but with a marginally deleterious effect in the trial overall.

Our framework for understanding BCG efficacy is biologically plausible because the most favourable outcome following exposure is stable immune tolerance, and because the immunology of TB differs fundamentally between infants, children and adults (10). Infants, with a developing adaptive immune system and reduced capacity of antigen-presenting cells, develop a disease phenotype similar to that observed in HIV-1-infected individuals. This results in higher mortality and more frequent disseminated TB in infants, but is ameliorated by BCG priming trained immunity and enhancing early *Mtb* containment (44). Ages 5 to 14 years represent an epidemiological paradox of low TB risk despite high *Mtb* exposure in high-burden settings (45). During this pre-adolescent period, boosting natural immunity to TB through vaccination may be less important, although BCG also protects against respiratory viruses, possibly through “trained” innate immunity (46). Differing from classical T and B cell immune memory, trained immunity refers to epigenetic modulation of innate immune cells; in the case of BCG, monocyte, NK and γδT-cells demonstrate enhanced IFNγ, TNF, IL-6 and IL-1β production though DNA chromatin relaxation at these gene promoters. From around age 15, TB risk dramatically increases and the phenotype of disease changes to classic pulmonary cavitary disease. If by 15 to 20 years post-vaccination acquired T cell protection wanes but trained innate immunity persists, then the host will have lost the beneficial BCG-related effects of T-cell-mediated immunity, but acquired a persistent hyper-reactive innate response. Such a response is seen in the transcriptional signatures predictive of TB risk, and may be responsible for the liquefactive necrosis that leads to clinical symptoms, organ damage and transmission in adults (47). Therefore, because TB disease is immunologically mediated, it is plausible that induced T cell sensitisation to a broad range of *Mtb* antigens could protect against the first encounter with the organism during an age of suboptimal immunity. However, it may have a reduced or adverse effect with repeated exposure in adulthood, when the immunological response is vastly different to that of infants, for whom BCG is clearly effective.

Past reviews have proposed that between-trial heterogeneity may be partially explained by latitudinal gradient in non-tuberculous mycobacteria (NTMs) exposure, particularly in studies employing one-stage TST testing (4). These arguments require that: *1)* NTM sensitisation decreases with latitude, *2)* TST positivity detectable only with two-stage testing frequently represents sensitisation to NTMs rather than *Mtb, 3)* NTM sensitisation confers immunity to *Mtb*, and *4)* because of NTM-conferred immunity to *Mtb*, BCG vaccination boosts immunity to *Mtb* to a lesser extent in those with NTM sensitisation than those without. Although point *1)* is well-established, latitudinal gradient also applies for *Mtb*. To our knowledge, evidence is conflicting for point *2)* (36) and derived from animal models for points *3)* (48) and *4)* (49). The greater rates of TB observed in NTM-naïve persons at an equivalent TST response (50) likely reflect a greater probability of *Mtb* infection. Re-analysis of the Chengalpattu study suggested somewhat lower protection in NTM-sensitised participants (51), although the statistical significance of this finding was unclear, while the one trial conducted across sites at multiple latitudes found similar effects by location (30). Although we do not discount the importance of NTM sensitisation, a greater effect of vaccination on clearly TST-negative participants and young children would also support our hypothesis. That is, if the lower efficacy of BCG distant from vaccination is attributable to accumulated exposure to *Mtb*, then exposure to both *Mtb* and NTMs could have similar effects in mitigating vaccine efficacy. As such, the recent success of the M72/AS01_E_ trial in *Mtb*-exposed, predominantly BCG-vaccinated adults in a high-burden setting, supports the need for revaccination with new antigens to mitigate any increased risk from past BCG vaccination (52).

We therefore believe that our hypothesis is supported by the evidence to a considerably greater extent than previously accepted explanations. However, as is common in empiric research, not every aspect of the analysis is perfectly consistent with the hypothesis. For example, our hypothesis does not explain the slight adverse effect suggested in the Georgia schools trial, although the very small number of cases is consistent with random chance and heterogeneity was not observed in this trial category. We also expected more TB cases in the vaccinated group late from enrolment in the Chengalpattu trial, which was seen to only a minor extent. As previously suggested, this may be explained by high rates of pre-existing *Mtb* exposure in both the control and vaccinated participants because of one-stage TST testing.

We propose that BCG vaccination protects against early post-vaccination exposure to Mtb, but is less effective or may increase rates of disease with later exposure, that the results of most or all past trials of BCG vaccination are consistent with this hypothesis and that this framework is more plausible than previously proposed explanations. We believe this explanation is also highly intuitive in retrospect, but may not have been recognised previously because Mtb sensitisation at recruitment has generally been excluded in trial participants based on TST. This phenomenon is infrequently observed because it is only seen in studies with long follow-up undertaken in high-burden settings, which are also the most logistically challenging to perform. Given that no clinical trial has found a statistically significant increased risk of disease, the increased rates of TB from late post-vaccination Mtb exposure may or may not represent an increased lifetime risk in high-burden settings. However, deferring episodes of disease could still have extremely important epidemiological effects, given that paediatric TB more often results in serious sequelae, whereas adult TB is more infectious and critical to perpetuating the epidemic. It is essential that future studies of TB risk and the effect of BCG vaccination present results disaggregated by age and time from exposure to fully elucidate these distinct reactivation profiles.

## Data Availability

As indicated in the manuscript, all code and new data are available at the link below.

https://github.com/jtrauer/bcg_tb_context_review

## Acknowledgements

We express our great thanks to the staff of the National Institute for Research in Tuberculosis for providing the disaggregated results used to generate the last panel of **Figure 3**, including Dr Srikanth Tripathy, Basilea Watson and others. We also gratefully acknowledge the contribution of the following individuals who assisted with data extraction for non-English language papers: Eike Steinig, Maddalena Cerrone and Joanna Zając.

