## Supplemental appendix for "Timing of *M. tuberculosis* exposure explains variation in BCG effectiveness: A systematic review and meta-analysis"

### Supplemental Appendix to Timing of *Mycobacterium tuberculosis* exposure explains variation in BCG effectiveness: A systematic review

November 23, 2020

**11 Recent excluded trial, H4:IC31 trial** **29**

**12 Full search strategy** **29**

### 1 Background

#### 1.1 Historical perspective

The bacillus of Calmette and Guérin was derived at the Institut Pasteur by serial passages of an isolate of Mycobacterium bovis through a medium of ox bile and was first given to humans orally in 1921.[23, 20, 42] However, from the earliest days of the study of BCG, opinions as to the efficacy and role of the vaccine have differed. In Europe, the charismatic Leon Calmette believed the vaccine to be so efficacious that controlled studies were unnecessary. This early enthusiasm was curbed by the Lübeck disaster in northern Germany around 1930 in which 77 infants died, although this was ultimately attributed to laboratory error and contamination or replacement of BCG with virulent *Mtb* organisms.[14, 23, 20, 50]

Formal clinical trials began from the 1930s with positive results reported from several early studies.[63, 23, 20] Towards the end of the first half of the $20^{th}$ century vaccination policies differed across continents, with countries of Scandinavia, South America, Japan and regions of continental Europe the earliest adopters.[6, 42] However, there was less enthusiasm for wide-scale vaccination in the UK and the USA, and large trials were initiated in these countries and in India to improve the evidence base to support BCG vaccination programs.[20, 42, 41] Features of these studies that were considered particularly important included assigning some participants to a control intervention, large numbers of participants, linkage of follow-up to existing surveillance systems and follow-up of those persons screened but excluded from the study (typically TST-positive persons).[41]

In the USA these trials were sponsored by the Public Health Service and showed only 30% protection, leading to recommendations for vaccination only as an adjunctive intervention in specific risk groups[46, 42, 7] (for reasons described below). Dramatic reductions in rates of TB disease, TB mortality and *Mtb* infection in the USA meant that older adults with LTBI had become the major epidemiological contributor to disease, while the elderly and those with existing LTBI were rarely considered for BCG vaccination.[3]

In the immediate post-war period, hundreds of thousands of children were immunised with the support of the Danish Red Cross Society.[33] This campaign was soon expanded beyond Europe, with international health and relief agencies responding to the considerable international TB epidemic with the International Tuberculosis Campaign,[20] through which 100 million people were immunised by 1958.[7] Part of the rationale for scaling up these programs was the feasibility of rolling out vaccination in settings in which case management-based interventions might not be feasible.

The WHO and UNICEF estimate that global coverage of BCG vaccination increased from 15 to 81% between 1980 and 1990.[62]

Ecological trends in TB burden resulting from BCG vaccination are difficult to study because of the broad range of epidemiological changes simultaneously influencing TB dynamics in any setting. In particular, universal BCG vaccination programs have typically been instituted in the context of general improvements in living conditions and other programmatic responses to TB. Nevertheless, broad ecological comparisons have been made across low-burden settings with markedly different policies, and an age-specific comparison between the universal adoption of vaccination in Scandinavia and the limited use in the USA suggests some effect of BCG vaccination on population disease burden in settings where burden is already declining.[13]

#### 1.2 Arguments against BCG

Common arguments against the widespread use of BCG vaccination have included the loss of the TST as a diagnostic test, local adverse effects from the vaccine, over-estimation of the risk associated with primary infection and trial endpoints that over-emphasise self-limited radiological changes associated with primary infection.[3, 42] Concerns that protection against infection in childhood might increase the rate of new infections among adults were alleviated by studies showing that the response to primary infection often ran a benign course in adults, as it did in children.[3] Trials in the USA with modest results and in populations with high prevalence of LTBI led to estimates that BCG could only prevent around 30% of cases among around 25% of the infection-naïve population and so could avert less than one tenth of all disease in a population.[42]

#### 1.3 Heterogeneity in findings

The marked heterogeneity in study results for the efficacy of BCG vaccination has long been recognised and cannot be explained by chance alone.[2, 23] Several hypotheses have been proposed for this variation in efficacy, although none has yet been accepted as the main explanation. The extent of heterogeneity has led to severe criticisms of attempts[15] to estimate a single value for the effect size of BCG vaccination. For example, P. E. M. Fine described the conclusion that BCG confers a 50% reduced risk of TB on average as "improper statistically and misleading for the immunological and public health communities, as it implies that variability observed is attributable to chance variations in study results. The implied logic is comparable to calculating the mean of the per capita incomes of Burkina Faso and of Switzerland and concluding that the world is, on average, middle class."[23] The finding of a negative efficacy or adverse effect has been previously but infrequently acknowledged.[56, 21]

#### 1.4 Systematic reviews

##### 1.4.1 Colditz, 1994

One of the most widely cited reviews of BCG efficacy was published in 1994 and reviewed 14 trials and 12 case-control studies.[15] The review found around 50% protection against pulmonary TB with both study types, and higher levels of protection against disseminated disease, meningitis and death. Co-variates were explored through regression models, with geographic latitude and study validity explaining around two-thirds of between-study variance, while mean age at vaccination, study duration and other co-variates did not increase the proportion of variance explained further.

##### 1.4.2 Sterne, 1998

A 1998 systematic review looked specifically at the question of change in efficacy of BCG vaccination with time and restricted inclusion to studies that presented results disaggregated by time from vaccination.[56] Nearly all analyses showed high degrees of heterogeneity, with seven trials finding a waning of efficacy with time and three showing an increase. Variation in efficacy at several time points was significant, but variation in efficacy more than ten years from vaccination was not, although this may simply reflect fewer data being available with longer time from vaccination. The estimate for protection more than ten years from vaccination, for which heterogeneity was modest, was 14% (95%CI -9% to 32%). Despite the heterogeneity, an estimate for the rate of waning immunity of 5% to 14% was proposed and no correlation between this rate of waning and the overall efficacy of the study was found. Of note, this review was published before the publication of the longest follow-up period of a clinical trial, being the 2004 paper on the 60 year follow-up of the native American vaccination study.[10]

##### 1.4.3 Abubakar, 2013

A systematic review of the duration of protection of BCG vaccination included both trials and observational studies.[2] The authors noted that estimates varied considerably, with the UK MRC trial finding a rate ratio of 0.22, while the Chengalpattu trial found a ratio of 1.05. Variation was considerable with latitude, while there was little variation with form of disease, study design or vaccine strain. Efficacy was significant during ten years of follow-up, while most studies did not present data or had too few cases accruing after 15 years of follow-up. Only five studies (one RCT and four observational studies) found a measurable protective effect after 15 years of follow-up, although efficacy declined with time and was greater in latitudes further from the equator and following stringent TST testing. Little or no protection was found for persons already infected *Mtb*.

##### 1.4.4 Mangtani, 2014

Another systematic review and meta-analysis from the same institution as Abubakar, 2013 aimed to explore associations with the variation in outcomes from 18 randomised or quasi-randomised clinical trials.[34] Absence of sensitisation to *Mtb* or non-tuberculous mycobacteria was suggested as a likely effect modifier, because of the stronger protection in children who had been stringently TST tested to exclude prior exposure and in studies further from the equator. Results were presented with disaggregation by age at vaccination and by whether two-stage ("stringent") TST testing was performed, along with disaggregation by geographic latitude. Age and latitude were able to explain all of the between-trial heterogeneity, although the independent effect of latitude even after inclusion of TST testing stringency may argue against NTM exposure being the mechanism underpinning the association between latitude and efficacy.

### 2 Supplemental methods

#### 2.1 General approach

The systematic reviews of Abubakar in 2013[2] and Mangtani in 2014[34] had many authors in common, employed the same methods and reported the same number of articles extracted. We employed identical methods of those used by this group to extend the review from 2009 to 31$^{st}$ December 2018. Our methods were also consistent with our registered protocol, available at CRD42019119676. Details of our search strategy are shown in Section 7 of this Supplement. The number of articles identified and their inclusion/exclusion in this review is presented in the study flow diagram. We focused on trials comparing BCG vaccination against control participants, as the highest forms of evidence available and present the following detailed narrative review of the results of these trials.

We also reviewed all cohort studies, as a secondary level of evidence, but did not include the other study types included in the previous reviews (classified as case-control, case-population, observational and outbreak studies by the previous review).

All studies published post-2009 and so not included in the previous systematic review by Abubakar et al. were reviewed by two authors (AK and BW). Note that this only applies to cohort studies and does not include any of the trials, as no additional eligible trials post-2009 were identified through our search strategy.

#### 2.2 Efficacy estimates

In determining the efficacy estimates of both the trials and the cohort studies, we used the following approach:

- If the end date of follow-up was not stated (as was the case in a minority of studies), this was assumed to be one year prior to the date that the article was submitted for publication
- If the end date of recruitment was not stated, this was assumed to be the end date of the follow-up period (which appeared to implied by several included studies)
- Zero counts were inflated by $\frac{1}{2}$ (except in the case of the Chicago mental health patients trial, because this study observed only one case of unconfirmed TB and we did not estimate efficacy for this trial)
- We recalculated all effect size estimates using standard formulas, which were similar to the estimates reported by the authors and in the prior systematic review for most or all trials
- Where person-years were reported by the study authors, these were used as denominator for hazard calculations
- Where person-years were not reported, person-years of follow-up were calculated using the total number of persons under observation at the mid-point of each study follow-up period
- Where the necessary data on follow-up were unavailable and only starting populations were available, we assumed that all participants remained under observation until the development of TB. The average follow-up period was therefore calculated as the total time from the end of recruitment to the end of follow-up plus half of the recruitment period. The cohort size was calculated as the starting cohort size minus half of the number of cases developing in that cohort, to account for failures (in the survival analysis sense).

#### 2.3 Classification

We applied our own novel approach to grouping the included studies into categories, differing from that used by Mangtani, who classified the studies as neonatal, school age and other age, with further classification according to TST testing. We did this because we consider the neonatal, school age and other age classification to be misleading, because many studies classified as other age predominantly recruited children. We also note that the Agra study recruited during the pre-school age of zero to five years.

We also consider that the classification of studies as either stringent or non-stringent TST testing may have oversimplified the process of testing prior to recruitment and that the terms zero, one-stage and two-stage are more informative.

Most importantly, two large trials were misclassified in the Mangtani review, with the Puerto Rico children study being incorrectly classified as non-stringent, when most participants received two-stage testing, and the Madanapalle study being incorrectly classified as stringent, when most participants received one-stage testing. These mistakes were reviewed blinded by three study authors (JMT, AK, RR) who were in agreement with the revised assessment. Each of these studies contributes the majority of the person-years of follow-up for the sub-group into which they were incorrectly classified.

Although the Mangtani et al. review reported its findings as applying to "protection against pulmonary TB", studies varied as to whether they reported on all cases of TB, pulmonary TB, bacteriologically-confirmed TB, culture-confirmed TB or radiologically diagnosed (and so presumably pulmonary) TB. We therefore present the case definition used by each study, or report "TB" where the trial does not specify further or only uses the terminology "TB" or "TB case".

We summarise the differences between our findings and those of Mangtani et al. as follows. Major differences between the results of our review and that of Mangtani et al. are:

- We propose an alternative approach to study classification, because we believe the neonatal, school age and other age classification is misleading
- We reclassified the Puerto Rico children study as two-stage
- We reclassified the Madanapalle community study as one-stage
- We report explicitly for each study on the case definition used where

provided or indicate whether the study applies to all forms of TB or a sub-group to the extent possible

Minor differences between the results of our review and that of Mangtani et al. are:

- We do not consider the Agra study to be a school age study, because we would understand that the age classification refers to the age of participants at trial entry and the participants in this trial were aged zero to five at entry and then followed for five years
- We assessed that the Georgia School study reported five cases of TB in the vaccinated, rather than the four assessed by Mangtani et al.
- As we recalculated all person-year estimates of follow-up, these values differ in many cases, almost always to a minor extent (except for one cohort study noted in the Table below)

#### 2.4 Risk of bias

We reviewed the risk of bias assessments presented by Abubakar et al. in Appendix 4 of their 2013 review [2] according to standard quality domains for clinical trials. Our assessment did not differ from that of the earlier review in any of these domains. However, we considered that the epidemiological context for TB and the diagnostic approaches used were of particular relevance and so also extracted this information, which we presented as the following extended narrative review and briefly in Table 2 (main manuscript).

### 3 Neonatal trials

#### 3.1 Saskatchewan native infants, Canada

A cluster randomised trial of BCG vaccination was undertaken in Qu'Appelle in Saskatchewan from 1933 to 1945.[22] Assignment was by family, with 306 and 303 allocated to vaccination and control respectively. The large majority received parenteral vaccination, with 21 participants being vaccinated orally. Cases of subsequent disease were diagnosed predominantly with x-ray. Follow-up extended to 1947, with the average follow-up duration being 6.6 years and 6.1 years for the vaccinated and controls respectively, such that virtually no children would have been observed into to their adolescent years. Rates of TB and TB-related mortality were found to be approximately five-fold lower in the vaccinated group, with six and 29 cases of TB, and two and nine TB-related deaths reported the vaccinated and controls respectively. No cases of TB were observed from 11 years of follow-up/age onwards, and all but two cases occurred in the first eight years of follow-up.

This period followed a TB epidemic in the area that reached extreme proportions around the 1880s, and although TB-related mortality had fallen approximately ten-fold over the four decades to 1927, rates of infection were likely still considerable during the study period. The proportion of control participants infected reached 68.7% by nine years of age, also implying continuing high rates of infection.

#### 3.2 Native infants, USA

In this study, 123 Native American infants were assigned to vaccination and 139 to control and followed with TST and x-ray, with three publications reporting on six to eight years of follow-up.[6, 4, 5] Recruitment took place from December 1938 to December 1940 through hospitals of the Turtle Mountain Agency in North Dakota and the Rosebud Agency in South Dakota. Annual radiological and TST assessment were performed as follow-up. Radiological evidence of TB was found in four of the vaccinated children and in 11 controls, while no TB-related deaths occurred in vaccinated infants and four in the controls.

For a discussion of the background burden of TB, see the description of the "native Americans in four states, USA" study below.

#### 3.3 New York infants, USA

This trial predominantly considered parenteral vaccination, but included some orally vaccinated infants in the first two years of the trial.[32] Infant participants from families in which a case of smear-positive or smear-negative pulmonary TB had occurred ("tuberculous families") were assigned to vaccination or control arbitrarily based on physician preference from 1926 to 1933, but alternate assignment was used from $1^{st}$ January 1933 onwards. Most children were enrolled in the first month of life and all within the first year of life, with TST (0.1 to 10mg old tuberculin) required if enrolment occurred after one month of age. Controls were compared against children whose TST converted from negative to positive following BCG vaccination. Active follow-up with clinical review monthly by nurses and 3-6 monthly by paediatricians was undertaken, with TSTs and x-rays at unspecified intervals. Despite extensive attempts at follow-up, fewer of the vaccinated than the control children were lost to follow-up (11 versus 31) in the early follow-up period.[32] The diagnosis of a TB-related death was confirmed were possible with autopsies, including microbiological investigations.

A first report in 1938 describes six and four deaths among 383 and 361 vaccinated and control children respectively.[30] The second report describes eight deaths in each of the two study groups, which then included 566 and 528 children in the vaccinated and control groups respectively.[31] By the time of the third report 1,005 vaccinated and 1,069 controls had been followed for up to six years, although the paper describes the outcomes of 463 and 476 vaccinated and control children respectively who had been exposed to tuberculous infection.[30] In this sub-group, five and six TB deaths accrued in the vaccinated and control children. This relative lack of protection was surprising, given that BCG vaccination had appeared highly protective against TB death (3 versus 18 deaths) in the arbitrary assignment period. Lower rates of radiological changes are mentioned in the vaccinated children, but no overall estimates of morbidity rates are provided.

TB burden in New York City declined approximately four-fold from 1900 to 1930, although TB-specific mortality rates remained as high as 300 per 100,000 per year.[29] Of the 476 unvaccinated children included in the trial, approximately one quarter (93) converted to a positive TST during the observation period.

#### 3.4 Chicago hospital-delivered infants, USA

Following x-ray screening of the whole family or the mother to exclude active TB, alternate newborns delivered at the Cook County Hospital in Chicago, USA were assigned to BCG vaccination or control, administered within the first week of life.[50] These infants were predominantly black and lived in disadvantaged areas of Chicago. Assignment began as alternate, but then changed to a 2:1 ratio and the children were then followed quarterly with clinical review, and six monthly with radiography and TST.

The first publication arising from this study reports three and 23 cases of TB and one and four TB-related deaths.[50] A second publication reports on the same follow-up period and focuses on the x-ray findings in 1,417 vaccinated and 1,414 control participants respectively, with 5,627 and 6,032 person-years of follow-up.[39] This paper reports a total of 14 and 42 children with x-ray changes in the vaccinated and controls respectively, with most being hilar and the remaining classified as parenchymal, pleural effusion or calcification. The majority of these radiological changes resolved in the vaccinated group, while most persisted or calcified in the control group. A third publication in 1948[47] presents results for the same observation period as the second.[39] Among newborns without known exposure to *Mtb*,

11 and 39 cases of TB had occurred in the vaccinated and unvaccinated respectively, while the number of deaths was one and seven respectively. A fourth publication, reports on 3,814 vaccinated and 3,014 control children (after vaccination had changed from a 1:1 to a 2:1 ratio) with up to 12 years of follow-up.[49] This publication reports 17 and 54 cases of TB and one and eight TB-related deaths in the vaccinated and control groups respectively, for a vaccine efficacy of 75% and 89% for TB disease and TB-related mortality respectively. A fifth publication reports on 5,426 vaccinated and 4,128 control children, finding 18 and 63 cases of TB and two and eight TB-related deaths in the vaccinated and control groups respectively.[53] The final publication reports on 23 years of follow-up, but for a smaller initial cohort, with results for 1,716 and 1,665 vaccinated and control participants respectively presented.[52] Presumably these results are the complete results for the subgroup of participants who remained in the study for this longer duration of time, as also reflected by the more balanced numbers during the initial alternate assignment period. In this smaller group, 17 and 65 cases of TB and one and six TB-related deaths had occurred in the vaccinated and control groups respectively. Despite this longer follow-up duration, case rates fell as the cohort was observed towards adolescence.

TB-mortality rates in the areas of Chicago from which these children came were estimated as 100 to 300 per 100,000 per year,[39] and 4% of children were TST-positive by 8 to 12 months of age and 30% were positive by $4\frac{1}{2}$ to five years.[49, 53] However, for the longer follow-up period, steep declines in TB burden would have occurred and nearly all cases of TB observed accrued in the first seven years of follow-up.[52]

#### 3.5 Chicago household contact infants, USA

Starting three years later than the hospital-delivered infants in 1940, a similar process was undertaken for children with significant home exposure to *Mtb*.[45] In most cases, vaccination was administered at seven to ten days of age, although if the mother was the index cases, this was deferred by up to three months and was further dependent on a negative TST to 1mg of old tuberculin. Alternate vaccination appears to have been practised for most of the enrolment period, although some of the reports have a moderate excess of vaccinated participants, suggesting that the early period of universal vaccination may have been included. The goal of the study was to determine whether vaccination was effective as a supplementary measure to excluding the infant from their mother.[50] Children were removed from their families for a variable duration depending on the nature of the contact (mother versus other household member) and clinical characteristics of the index patient's disease ("open" versus "closed"). They were reviewed in clinic every six months for anthropometry, TST and x-ray.

The first publication from this trial reports one and four cases of TB, with zero and three TB-related deaths, occurring in 58 vaccinated and 63 control participants respectively.[50] The second publication reports on up to seven years of follow-up of a cohort of unclear size, by which time two and five TB cases, with one and four deaths, had occurred in the vaccinated and control groups respectively.[47] By the third publication, there were 139 vaccinated and 128 controls, among whom three and six cases of TB, with one and four deaths, had occurred in the vaccinated and control groups respectively.[45] The fourth publication describes results after up to 12 years of follow-up of 276 vaccinated and 218 control participants, at which time four and ten cases of TB, with one and four TB-related deaths, had occurred in the vaccinated and control groups respectively.[49] By the fifth report which described up to 15 years of follow-up, 311 vaccinated and 250 controls had been enrolled, among which the same number of TB cases and deaths had occurred as in the fourth report.[53] The last report describes follow-up for up to 13 years of 231 vaccinated and 220 controls (although the explanation for these lower numbers is unclear), with an average follow-up duration of $5\frac{1}{2}$ years for the vaccinated and six years for the controls. By this time, one further case of TB had occurred in the controls, for final case numbers of three and eleven in the vaccinated and controls respectively, with zero and four TB-related deaths.[48] The radiological changes found were noted to be less marked in the participants who had been vaccinated in several of these reports, despite the small numbers.

Very high rates of infection in this setting were evident from the high rate of TST conversion in the controls, with more than half (57.6%) of the controls becoming TST-positive by 5 and a half years of age. The TB-specific mortality rate in the communities in which the trial was undertaken was estimated at 330 per 100,000 per year.

#### 3.6 Mumbai infants, India

A trial in a paediatric clinic in Mumbai enrolled 696 infants, of whom 396 were randomly assigned to vaccination in the first week of life and 300 were assigned as controls.[38] 662 participants were followed with 6-monthly TST and clinical examination for the study duration of 30 months. X-rays and/or other tests were done in the case of clinical suspicion. In the vaccinated and unvaccinated group respectively, 22 and 27 cases of "primary TB" were diagnosed on the basis of TST results alone, although most of these had abnormal radiographic features (including all of the cases in the BCG vaccinated group). More cases of TB occurred in participants with close contact with a TB patient and with larger family size, leading the authors to speculate that intense exposure may have overwhelmed the effect of vaccination, although there was no evidence that exposure modified the effect of vaccination and case numbers were small.

High transmission is suggested by the 9% TST conversion at 24 months in the control group.

### 4 Other trials in young children

#### 4.1 Agra pre-school children, India

This trial in 2,930 slum-dwellers of Agra City in northern India recruited children aged zero to five years, screened participants with TST, and assigned half of the TST-negative (<10mm) children to BCG and half to control.[37] TB was diagnosed with radiological screening followed by sputum microscopy and culture for those with abnormal x-rays with follow-up extending for five years. Ten cases of TB occurred in the vaccinated and 25 in the unvaccinated, suggesting an overall vaccine efficacy of around 60%. The vaccine efficacy for children in a household in which a TB case did and did not occur can be estimated at 53% and 67% respectively.

High ARTIs are suggested by 8.1% and 58.0% of the children aged under five and of the total slum population respectively showing evidence of infection. The authors also report extreme rates of disease, with a prevalence of radiologically active TB for persons aged five and above of 5.0%.

#### 4.2 Chicago housing project, USA

In this study, TST-negative, predominantly black children from the Ida B. Wells federal housing project in inner southern Chicago aged up to twelve years were alternately vaccinated or left as controls.[47] TST was with 1mg old tuberculin and follow-up was with annual x-ray and TST.

The first publication from this study describes up to six years of follow-up of 699 and 625 vaccinated and control children, with zero and three cases of TB in each respective group and no TB-related deaths in either.[47] The second report describes up to ten years of follow-up of 777 vaccinated children and 805 TST-negative controls, with no additional cases or deaths accruing.[49] The third publication reports on a greater number of children, although the number alternately assigned is not reported. No further cases of TB or TB-related deaths had occurred by the time of this report.[53]

### 5 Paediatric trial with short follow-up

#### 5.1 Jeremie community, Haiti

The purpose of this cluster randomised controlled trial was to determine whether isoniazid-resistant BCG could be of use in maintaining the effect of vaccination, regardless of whether isoniazid preventive therapy was being concurrently administered.[61] In addition to trial arms providing a potent strain of BCG and an isoniazid-resistant strain derived from the same strain, a placebo group was also included in a ratio of 2:2:1, with half of each of these three groups provided with isoniazid preventive therapy for 8 to 10 weeks. Vaccination was administered to participants with a <6mm response to 5 TU TST in a low socio-economic society of Jeremie in South-West Haiti from November 1965 to June 1966. Because TST-positive persons were excluded and around three-quarters of the population were TST-positive, participants were predominantly aged under 20. Furthermore, the low yield of negative TST results in the over 20s led to their exclusion from the later phases of recruitment.

Participants were followed up with chest x-ray at baseline and annually thereafter, and TST was repeated after one year. The description of the study focuses on the immunological responses of participants across the vaccination and preventive therapy groups, although TB case rates to the end of three years of follow-up are also provided. At this time, ten bacteriologically-confirmed cases of TB had been identified among the 629 participants assigned to the unvaccinated control groups (regardless of isoniazid assignment), while eight bacteriologically confirmed cases had been identified among the 2,545 participants assigned to the two forms of BCG (regardless of isoniazid assignment). Of these, one case occurred in the 635 vaccinated with conventional BCG not treated with isoniazid and five occurred in the 338 vaccinated with placebo and not treated with isoniazid. Appropriately, these latter numbers are those that have been used in past meta-analyses.[34]

Initial screening found that TST positivity increased steadily with age in the study population, reaching 90% by the participants' late 20s. TB rates of 105 per 100,000 per year across the vaccinated groups and 530 per 100,000 per year across the placebo-assigned groups were observed, also consistent with a very high-burden setting.

### 6 Paediatric trial in high-burden setting with longer follow-up

#### 6.1 Puerto Rico children, USA

An island-wide study of BCG vaccination was planned in Puerto Rico and 96% of potentially eligible children were screened for entry from 1949 to 1951.[42, 41, 16] For school-aged children, TST screening was with 1 and 10 TU of PPD-RT-19-20-21, while pre-school-aged children received only a single 10 TU dose. A third dose of 100 TU was administered, although this did not affect enrolment. This study has previously been incorrectly classified as having screened participants with non-stringent TST.[34] Despite aiming to recruit most of the island's children, ultimately only 191,827 of 1,088,600 children in the island entered the observation cohort, because of insufficient community acceptance, of which 82,269 were excluded for a positive TST response and the remaining 109,558 were offered trial enrolment. In successive age brackets of three years, the middle year was assigned to control and years one and three were vaccinated. After a further 31,586 participants then refused randomisation, 50,634 were allocated to vaccination and 27,338 to control, reflecting the 2:1 assignment ratio.

Follow-up was through established surveillance systems, with the large majority of cases reported through Puerto Rico's twenty TB centres distributed throughout the island. There was no obvious difference in the organ manifestation of the resulting TB disease. Although children aged one to 18 were targeted, ages 8 to 14 had the greatest representation and half of the study population were aged 7 to 12 because of the age profile of consenting children and exclusions for TST positivity. Therefore, this study predominantly considers protection against the high-risk adolescent period in a high transmission setting.

Although case rates fluctuated and declined somewhat over the follow-up period of the trial, they remained modestly lower in the vaccinated than the unvaccinated group during each 5-year period of observation. Specifically, the first of two reports on this trial describes 93 and 73 TB cases occurring in the vaccinated and control groups.[42] The latter report in 1974 describes 141 and 186 TB cases in the respective groups.[16]

Puerto Rico in the mid-20$^{th}$ century was densely populated and had a very high rate TB disease. An annual risk of infection of 4% or greater is suggested by 21% of six year-olds and 69% of 18 year-olds being TST-positive at screening. The TB-specific mortality rate peaked at 332 per 100,000 per year in 1933 before falling to 179 and 33 per 100,000 per year in 1948 and

1955 respectively, with rates approximately sixfold that in the continental USA.[42]

### 7 Paediatric/adolescent trials in declining burden setting with longer follow-up

#### 7.1 Native Americans in four states, USA

A controlled BCG vaccination study was initiated from 1935 to 1938 in around 3,000 Native Americans aged one month to 19 years, with follow-up facilitated by provision of health care primarily through the Indian Health Service.[7, 55] Participants negative to two-stage TST (at doses of 0.00002 and 0.005 mg PPD, cut-off for positivity not stated) were alternately assigned to vaccination or control and actively followed until 1947 with radiographic and TST assessments (annually until 1944).

The earlier publications reporting on this trial focus on the radiological findings. Although this approach was initially criticised for not focusing on clinical cases,[3] an effect on mortality was observed and follow-up was later extended (as described below). The first report describes three years of follow-up, during which six and 59 participants developed tuberculous lesions in the vaccinated and control groups respectively.[57] The second paper reports on six years of follow-up, during which 40 and 185 cases of TB and four and 28 TB-related deaths had occurred in the vaccinated and controls respectively.[9] Two reports in 1948 describe up to eleven years of follow-up, finding a range of radiological changes possibly attributable to TB (including calcified nodules and changes of doubtful aetiology) in 182 and 423 participants in the vaccinated and control groups respectively, along with six and 53 TB-related deaths.[5, 6] A fifth report describes the results obtained following a repeat visit to the study areas with attempts to determine causes of death of study participants 15 years after the initiation of the study.[8] By this time, a total of 12 and 65 deaths had accrued in the vaccinated and control participants respectively. Although the authors speculated that protection may have begun to wane after ten to eleven years, more TB-related deaths were observed in the control participants in most years even after this time. A subsequent publication soon after describes a reduction in the rates of a range of radiological manifestations of TB in the vaccinated, including "primary", "reinfection", pleural and miliary forms.[55] Rates of radiological evidence of TB increased slowly to the sixth year of follow-up and declined thereafter, likely reflecting the increased risk of disease towards adolescence. The last report of the $20^{th}$ century describes 18 to 20-years of follow-up to

1956 in which the mortality status of over 99% of participants was successfully determined, usually with bacteriological confirmation.[7] There were 13 and 68 deaths in the vaccinated and control groups respectively, for a TB-specific cumulative mortality of 0.84 and 4.7% during the period 1936 to 1956. More deaths continued to accrue in the control group until around 15 years of follow-up, with the greatest difference in TB-specific mortality being in those aged 10 to 24 years, while in those aged 25 to 34 a total of only three deaths occurred in all participants.

The final report describes a careful attempt to locate information on participant outcomes including TB disease from 1948 to 1998, after active case finding had ceased.[10] In 1,483 BCG-assigned participants followed past 1948, 36 cases of definite or probable TB occurred, while 66 occurred among the 1,309 assigned to placebo. The case rates over this time were 66 and 138 per 100,000 per year in the vaccinated and unvaccinated groups respectively, for a vaccine efficacy of 52% (95%CI, 27%-69%). Moreover, rates of TB were higher in the placebo group during each decade of follow-up from 1948 to 1998, although numbers of cases were small in the last two decades.

TB rates in Native American populations in the late $19^{th}$ and early $20^{th}$ centuries were extreme, with TB responsible for more than half of all deaths in the late $19^{th}$ century and incidence as high as 2,700 per 100,000 per year in the 1930s. The study was preceded by a TST survey which was consistent with ARTIs in of 5% or greater, with 90.0% of those aged 20 to 24 TST-positive.[7] However, the widespread use of antituberculous agents from the 1950s saw TB rates fall dramatically, with the study authors reporting a two to threefold decline in ARTI from 1937 to 1954 in younger age groups. Subsequently, case rates were decreasing by up to 40% per year and annual risk of infection by up to 12% per year through the 1960s, representing a dramatic interruption to transmission around this period.[44]

#### 7.2 Georgia schools, USA

A vaccination trial in schools was initiated in Muscogee County in Georgia, USA in 1947.[54, 41] Around half of children were TST-positive, with 4,839 children TST-negative and eligible for enrolment. Participants were almost all aged 5 to 19, with an average age of 11.4 years. Two-stage TST at doses of 5 and 100 TU PPD-RT19-20-21 was used to exclude a considerable proportion of children who were positive to the higher dose. The numbers assigned to vaccination and control were 2,498 and 2,341 respectively, with many of those assigned to vaccination re-tested with TST after six months and three years and re-vaccinated if they remained negative.

The first report on this study focuses on the TST responses after up to three years of follow-up.[54] The second publication reports on 12 years of follow-up, with 35 cases of TB occurring in the entire cohort, including the TST-positive. Of these, two cases each occurred in the TST-negatives assigned to either vaccination or control. By the third publication, 47 cases of definite tuberculosis had occurred across the cohort.[18] This publication defined "definite TB" as consistent x-ray findings or extrapulmonary manifestations, together with either bacteriological confirmation or a TST ≥10mm to 5 TU PPD. Of the definite cases, three and five cases of TB occurred in the vaccinated and controls respectively. Seven of these eight cases occurred in years six to 15 of follow-up, with none occurring in the first five years.

Of the controls, 7% showed TST responses of ≥5mm and the county was noted to have TB-specific mortality rates "somewhat below that of the rest of the country".[41]

#### 7.3 English cities, UK

From 1950 to 1952, around 56,700 school children without clinical TB at entry or known recent exposure to *Mtb* aged 14 to $15\frac{1}{2}$ years were enrolled in Birmingham, Manchester and north London.[60, 35] Those who were negative to two-stage TST (with 5 and 100 TU old tuberculin at <5mm) at enrolment were randomised "according to the final digit of their record card" to BCG, vole bacillus vaccination and no vaccination, with around 27,000 concurrently assigned to BCG or control (with vole bacillus not used in the London sites). All participants (including the TST-positive) were actively followed with a variable number of clinical, TST and radiological assessments around the time of finishing school, as well as with the same assessments every 14 months thereafter. The main comparisons were made between each vaccinated group and its concurrently enrolled controls.

The first publication reported considerably lower rates of disease in the BCG-vaccinated group than the unvaccinated over the first $2\frac{1}{2}$ years of follow-up.[60]. This was again observed in the second publication, which provided complete data on the first five years of follow-up and reported 27 and 151 definite cases of TB in the vaccinated and control groups respectively,[35] the third publication, which provided complete data on the first $7\frac{1}{2}$ years of follow-up and reported 48 and 213 definite cases in the vaccinated and control groups respectively,[1] and the fourth publication, which presented complete results to the end of 15 years of follow-up and reported 56 and 240 definite cases in the vaccinated and control groups respectively.[36] Rates in the previously TST-sensitised group were intermediate between those of the vaccinated and control groups (who were TST-negative at enrolment). BCG appeared particularly protective against severe forms of TB (i.e. CNS and miliary) and a trend towards lower rates of hilar lymphadenopathy and less extensive disease among participants developing pulmonary forms of TB was suggested.

The final publication arising from this study reported on 20 years of follow-up of the participants, with only 27 cases of TB developing between 15 and 20 years.[28] Of these, six and five cases of TB occurred in those (initially TST-negative) participants randomised to BCG vaccination and control respectively. The rate of disease was lower in the BCG-vaccinated group than the unvaccinated group in each $2\frac{1}{2}$ year interval until $12\frac{1}{2}$ years of follow-up and comparable in the last three $2\frac{1}{2}$ year intervals. Over the 20 year follow-up period, the protective efficacy of BCG vaccination was 77%, with efficacy estimates of 84%, 69%, 59% and -9% calculated for each 5-year period from recruitment to 20 years of follow-up. Although the latter estimates were associated with wide confidence intervals due to the steep decline in the number of TB cases occurring, the trend towards lower efficacy was considered statistically significant.

These results of this study imply considerable protective efficacy for somewhat over ten years from vaccination, which waned thereafter in a setting of declining TB burden. The results of this study led to changes in vaccination policy in the UK, with vaccination for 10 to 13 year-olds introduced from 1953,[28] but not in the USA where a different strain had been provided across a broader range of ages.[23]

Around 40% of children were TST-positive at screening, suggesting an ARTI of >3% prior to the study commencing, with well-documented rapid declines in TB burden thereafter. Tuberculosis mortality in England and Wales fell from 19,721 in 1949 to 1,840 in 1969, as effective chemotherapy (with streptomycin and para-aminosalicylic acid) and preventive measures (such as the eradication of bovine TB) were implemented.[36] This steep decline was also reflected in the number of TB cases occurring in the study, with 227 cases occurring in the TST-negative control group in the first ten years of follow-up but 21 occurring in the latter ten years. Similarly, 519 and 91 cases occurred in the study as a whole over the earlier and later ten year periods, although these numbers would be slightly affected by cohort attrition.[28]

### 8 Adult trials with short follow-up

#### 8.1 Chicago mental health patients, USA

In a mental institution in Chicago, most of the 4,500 patients aged up to 66 years assessed were positive to two-stage TST performed at an interval of seven months with 1mg old tuberculin.[47] Of the remaining 35, 20 were vaccinated. After four years of follow-up, no cases of TB had occurred in the vaccinated and there had been one possible case (described as "bilateral minimal arrested tuberculosis") in the controls.

#### 8.2 Chicago nursing students, USA

From 1940 to 1953, nursing students of Cook County Hospital in Chicago, USA who were negative to two-stage TST testing (2 and 10 TU of old tuberculin with <6mm cut-off) were alternately assigned to BCG vaccination or control.[46, 47] The policy on revaccination following negative TST results changed during this period, and so during the subsequent years (1953 to 1961) newly enrolled nurses were still followed, although vaccination was extended to all new enrolments during this interval, with this period not considered part of the trial. TST-negative nurses were not permitted to work on TB wards, which would be expected to bias this study towards a null result.

The first publication from this trial reports follow-up of 142 vaccinated and 199 control participants for up to seven years, in which zero and three cases of pulmonary TB occurred.[47] The second publication reports 231 and 263 vaccinated and control participants followed for up to 12 years, with two and five cases of TB occurring in the vaccinated and control groups respectively.[49] The third publication reports 269 vaccinated and 281 controls, with the same number of TB cases developing.[53] The final publication reports on 231 and 263 vaccinated and control participants, in which two and five cases of TB occurring in the two respective groups.[46] One of the two cases in the vaccinated group was non-pulmonary (cervical adenitis) and most of the remaining cases were described as being minimal. The period of follow-up appears to have been the duration of the nursing degree or three years, although one reported TB case in each group occurred after three years and four months.

Antituberculous chemotherapy was unavailable through most of the study period (streptomycin available from 1948, isoniazid from 1952) and despite their exclusion from TB wards, even the TST-negative nurses had high rates of TST conversion (57%, 79% and 89% converted to high-dose 100TU TST by one, two and three years respectively), suggesting high levels of exposure for all participants.

#### 8.3 Chicago medical students, USA

From 1939 to 1952, medical students (average age 23.3, range 20 to 37 years) were screened with TST using a first dose of 2 TU old tuberculin, followed by a second dose of either 10 or 100 TU or both.[51] Those with reactions of <6mm were then randomly assigned to BCG vaccination or placebo. Follow-up duration is not specified, but all 15 cases that developed TB (including the TST-positive at screening) are listed with a year of their studies in which the disease developed, implying that follow-up was for the duration of their studies, which has previously been assumed to be four years. Zero and three cases of TB occurred in 324 vaccinated and 298 controls medical students respectively. Most cases of TB that were identified were described as "minimal", although this group was predominantly comprised of the initially TST-positive.

As for the nursing students, substantial rates of TST conversion in the controls suggested high levels of exposure to *Mtb* in this setting (24% and 56% converted to high-dose 100 TU TST by one and two to three years respectively).

#### 8.4 Rand Mines, South Africa

A trial in South Africa recruited young adult males commencing work as miners.[14] For the first seven months of the study (1st October 1964 to 1st May 1965), all new entrants were vaccinated. For the following three years (until 1st February 1968), entrants were alternately assigned by company number to vaccination or control and followed until 30th April 1968. Screening at entry with chest x-ray was undertaken. However, Heaf testing was only performed for controls enrolled from June 1967 onwards and so did not appear to affect trial eligibility, such that the trial likely includes many participants who were TST-positive at baseline. This was the reason for the trial's exclusion from the 1998 review on duration of protection,[56] although it was included in the subsequent reviews by Mangtani and Abubakar. Follow-up was likely short, as the annual turn-over of miners was stated to be 90%, such that we assume that the average period of time that each individual miner would have been present in the mine would have been approximately 3.6 months.

The main outcome was active TB identified through the existing health system servicing the mine, which employed 6-monthly radiographic examination for all miners. Those developing TB in the days following entry were excluded and attempts were made to ensure no cases of TB were missed by review of chest x-rays performed as miners left the mine. BCG was found to be efficacious, with 54 cases occurring in the vaccinated and 98 in the unvaccinated. However, this estimate applies to all participants reported by the study and was not limited to those who were recruited during the alternate assignment period (or those initially TST-negative). Protection was also suggested in those alternately assigned, with 29 cases in the vaccinated and 45 cases in the unvaccinated, among 8,317 and 7,997 miners respectively.

Significant rates of infection in this population are suggested by around three-quarters of miners being Heaf grade one or greater and half being grade two or greater. This rate is lower than in some other studies of miners, which was thought to be attributable to a greater proportion of novice miners.

### 9 Adult trials in declining burden settings with longer follow-up

#### 9.1 Muscogee-Russell Counties community, USA

These two contiguous counties of Georgia and Alabama had a combined population of 155,000 in 1950, with one quarter of the population rural and one third African-American.[41, 42] All residents of the counties aged 5 and above were eligible to participate and were screened with radiography to exclude active TB and one-stage TST (5 TU PPD-RT-19-20-21, cut-off <5m), after which alternate birth years were BCG vaccinated. By contrast to the paediatric trial in Puerto Rico, outreach and community engagement was more successful and about half the population participated, with higher participation rates in younger ages. Just over half of these participants were TST-negative and so allocated to vaccination and control groups. Approximately 70% of the population was white, and (after a greater number of adults had been excluded for positive TSTs) 60% of participants were aged 5 to 19. As for the Puerto Rico trial, follow-up was through established surveillance systems, and continued for seven years.

The first report of this study describes the first seven years of follow-up and reports 17 and 28 cases of TB in the vaccinated and control participants respectively, with vaccine efficacy estimated at 36% (which was considered non-significant).[42] The second report describes 14 years of follow-up and reports 26 and 32 confirmed (by microbiological or autopsy findings) or presumptive cases of TB (positive TST at >10mm to 5 TU PPD with consistent chest x-ray findings or extrapulmonary clinical manifestations), including both pulmonary and extrapulmonary forms.[17] The last report of the trial describes 32 and 36 cases of confirmed or presumptive TB cases (as defined above) in the vaccinated and control participants respectively.[19]

The TB-related mortality at this time was 25 per 100,000 per year, which was comparable to that of the USA as a whole. Modest rates of transmission are also suggested by the higher rates of disease in TST positives than TST negatives, being the reverse of what had typically been observed in earlier trials, such as those undertaken in health care workers.

#### 9.2 Madanapalle community, India

A trial was undertaken in the small southern Indian town of of Madanapalle in Andhra Pradesh and its surrounding villages. The study involved mass screening with x-ray and TST testing from 1950 to 1955, with linkage to TB treatment, which had previously not been available. Two further radiological surveys were undertaken in 1957-58 and 1964-65. Participants with TST <5mm were randomly allocated to BCG or control and followed for up to 21 years, with around half the participants aged under 15 at entry.[26] Two different approaches to TST testing were undertaken during the course of the study, with a 1, 10, 100 TU regimen used initially, and those negative to the 1 and 10 doses included. This regimen was progressively replaced by positivity to a single dose of 5 TU being the inclusion criterion (who were also subsequently also given a 100 TU dose, although this test did not affect trial enrolment, as in the 1, 10, 100 TU regimen).[27] The large majority of study participants (>95%) were tested with the 5, 100 TST regimen, making this study predominantly a one-stage TST study.

Assignment was by individual randomisation using cards, with a higher drop-out rate in those assigned to vaccination, such that 5,069 persons were assigned to vaccination and 5,808 to control. The second report of the trial found 11 and 29 cases in the vaccinated and control groups respectively.[27] The third report described 35 and 53 cases, with 18 and 31 of these being bacteriologically-confirmed respectively, giving a vaccine efficacy of 24% overall and 33% for bacteriologically-confirmed cases.[25] The final report provides results including presentations from the seven rounds of active screening and from passive case detection until 1971.[26] It focuses on smear-positive ("bacillary") cases, which is likely to explain the absence of any observable effect in children, with virtually no cases diagnosed before adolescence. It reports 33 cases of TB in the vaccinated and 47 in the unvaccinated participants, for a vaccine efficacy of 20%. Rates of TB were higher in the controls for the first nine years of the trial, but higher in the vaccinated from 16 to 21 years of follow-up. The authors concluded that vaccination may defer the timing of disease onset and that there was "some evidence that ... the vaccinated persons continue to produce cases when the corresponding 'controls' have ceased to do so."

The setting had a high burden of TB, with an annual risk of infection that was estimated by the study authors to be 2-4%, and a prevalence of smear-positive TB of around 570/100,000 at a preceding survey in Madanapalle Town. The vaccination study was linked to an "all out attack" on tuberculosis, with the recent availability of streptomycin and PAS, and isoniazid becoming available in the early stages of the study. TB mortality was estimated at $\geq$200 per 100,000 per year prior to the study, but fell approximately tenfold over the first four rounds of screening to 21 per 100,000 per year. Although an equivalent fall in TB prevalence was not apparent, a shift of cases towards older age groups was observed, which was likely attributable to longer survival of those with more advanced TB under treatment.[24] Moreover, the predominance of cases in the initially TST-positive (with rates approximately double in this group compared to those initially TST-negative and randomised to vaccination or control) suggests an epidemic that is substantially driven by late reactivation cases and is consistent with a declining epidemic.

### 10 Adult trials in high-burden setting with longer follow-up

#### 10.1 Lincoln State School, USA

In 1947, resident patients of a school in Illinois State (USA) for persons with intellectual disabilities of any severity were studied.[12] Of the 5,200 residents, 1,025 were included in the trial if TB was excluded and two-stage TST testing was negative. TSTs were at concentrations of 1:1000 and 1:100 old tuberculin, which is presumed to be 10 and 100 TU, with cut-off diameter for positivity not stated. 531 were assigned to vaccination and 494 to control, with 258 of those assigned to vaccination re-vaccinated on the basis of subsequent negative TST testing. Follow-up was facilitated by the low rates of discharge for the patients. Twelve years later, follow-up revealed twelve and eight cases of TB in the vaccinated and controls respectively, while for TB-related deaths the numbers were four and two. The clinical details for all but one of the vaccinated TB cases and all but two of the control cases imply pulmonary involvement, such that up to three included cases may have had extrapulmonary TB. Although the institution is referred to as a "school", a significant proportion of residents were adults, as evidenced by the ages of the reported cases. The mean age (presumably at trial entry) of persons developing TB was 20.7 in the vaccinated and 16.0 in the controls.

TB was known to be a particularly major problem in persons with mental illness in this setting. Rates declined steeply during the study period, with a reduction of case rates by 95% from 1945 to 1958. However, incidence remained higher in newly admitted patients and was many times higher than that in the general community.

#### 10.2 Chengalpattu community, India

The largest randomised controlled trial of BCG vaccination that included adults was undertaken in Chengalpattu, in Tamil Nadu in southern India, with the study area comprising 209 villages and one town, approximately 40km west of Chennai.[59, 11] BCG vaccination had not been previously provided in Chengalpattu, which was a predominantly rural setting with a very high burden of TB, particularly in males. The study included participants regardless of past exposure history, with the only exclusion being babies aged less than one month. A total of 265,172 individuals were identified from a population of 364,819, of whom around 115,000 were TST-negative at enrolment (with only those aged one year and above tested). TST positivity was defined as a reaction of ≥8mm based on the profile of responses observed in the study. The placebo control was a non-scarring preparation of dextran with a similar appearance to BCG, while the active treatments were two doses of two commonly used types of BCG (French seed lot 1173 P2 and Danish seed lot 1331). Patients were recruited from 1968 to 1971 and followed for 15 years until 1987. One third of participants were individually randomised to placebo and one third to each of the two doses of vaccine (0.1mg and 0.01mg), giving a 2:1 ratio of vaccinated to controls. Those assigned to the vaccine groups were further factorially assigned to the two different vaccine strains in addition to the two doses. Participants aged ≥10 were screened for active TB at intake with chest x-ray followed by sputum smear and culture if TB was suspected radiographically (although the intervention was not modified based on initial TB status).

Follow-up was then undertaken with population-wide radiographic screening for those aged 5 and above at least every $2\frac{1}{2}$ years using similar diagnostic approaches to those at intake (with additional follow-up rounds in the first and fourth years). Further selective follow-up of high-risk individuals was also undertaken every ten months, along with continuous passive case finding. Two to three times between each resurvey, each village was visited and chest x-ray performed in those with symptoms or previously suspicious lesions, with those diagnosed with TB treated with antibiotics as outpatients. TB was diagnosed on the basis of a single positive culture result because smear microscopy was considered less reliable in this setting. Migration out of the study catchment was significant, with follow up of 89, 83, 78, 65 and 58% at 2.5, 5, 7.5, 10, 12.5 and 15 years respectively, although survival analysis techniques were employed to account for this.

The first report from this trial describes 124 and 47 cases of bacteriologically confirmed TB in the vaccinated and control participants respectively, with the vaccinated group being approximately double the size of the controls.[59, 11] A later report of the study in 1983 covers the first $12\frac{1}{2}$ years of follow-up and describes 192 and 93 cases of TB confirmed on at least one culture in the vaccinated and controls respectively (of which 125 and 65 were positive on two cultures).[58] This report also highlights the protective effect of the vaccination on children as they age towards adolescence in this high-burden setting, with a protective efficacy of 48% observed in those aged zero to 14 at entry in years five to $12\frac{1}{2}$ of follow-up. The first report on the full trial follow-up to 15 years gives slightly different cohort sizes, describing 117,718 participants TST-negative at entry and randomised, with 380 and 180 cases of TB developing in the vaccinated and control groups respectively. A subsequent report on the same follow-up period focuses on the effect of non-tuberculous mycobacterial infection.[43] Although this report suggests that there may have been a protective effect of BCG vaccination in those with absence of sensitisation to non-tuberculous mycobacteria (as evidenced by a negative response to PPD-B), the statistical significance of this finding was not clear.

The authors speculated that the study methods and BCG vaccine strains were unlikely to explain the negative results. This was affirmed by two WHO-organised workshops which did not find significant methodological flaws in the study's approach, and were unable to determine the reasons for the unexpected results.[23] They noted that by comparison to other trials, TB rates were particularly concentrated in those who were TST-positive at entry (>90% of cases), suggesting an important role for reactivation, in addition to primary progression, even in this highly endemic setting. They went on to speculate that BCG may protect against reactivation but not against exogenous reinfection, which was also likely significant in this setting given the moderate to high burden of TB. This was reflected by most study participants being TST-positive by age 24, although burden declined somewhat over the study period.

The study was originally intended to be one of two trials to distinguish between two alternative theories as to the reasons for the heterogeneity in observed efficacy, namely BCG strain differences and NTM sensitisation.[23] However, the companion trial in northern India was never initiated.

#### 11 Recent excluded trial, H4:IC31 trial

A recent South African trial of the novel vaccine H4:IC31 assigned 990 BCG-vaccinated, HIV-uninfected, IGRA-negative adolescents aged 12 to 17 to this novel subunit vaccine, BCG revaccination or placebo.[40] Participants were followed up for two years and the primary efficacy outcome was any positive QFT during this period. BCG vaccination protected against the secondary endpoint of sustained QFT conversion, but not against the primary endpoint. Rates of TB disease were not reported as a study endpoint, because the study's power was anticipated to be too small for this to be detected, and indeed no cases of active TB occurred in either group during the follow-up period (personal correspondence Hatherill).

#### 12 Full search strategy

We searched MEDLINE, MEDLINE in-process, BIOSIS, EMBASE, Cochrane Database of Systematic Reviews, ACP Journal Club, Database of Abstracts of Reviews of Effects, Cochrane Clinical Answers, the Cochrane Central Register of Controlled Trials (CENTRAL), the Cochrane Methodology Register, the Health Technology Assessment database, and the NHS Economic Evaluation Database, from 2010 to December 2018.

The following is the search strategy used in Ovid MEDLINE:

1. tb.tw (35998)
2. exp Tuberculosis/ (186821)
3. tuberculous.tw. (26930)
4. tuberculos$2.tw. (159515)
5. tubercular.tw. (3414)
6. phthisis.tw. (813)
7. tuberculoma$1.tw. (2381)
8. pott$ disease.tw. (703)
9. tuberculid$1.tw. (300)
10. scrofuloderma$1.tw. (185)

11. scrofula$1.tw. (132)
12. Mycobacterium bovis/ (12829)
13. Mycobacterium tuberculosis/ (46736)
14. tubercle bacill$.tw. (4312)
15. mycobacterium africanum.tw. (161)
16. mycobacterium microti.tw. (140)
17. mycobacterium canetti.tw. (3)
18. mycobacterium bovis.tw. (5934)
19. or/1-18 (244708)
20. BCG Vaccine/ (18550)
21. BCG.tw. (20644)
22. (bacill$ adj3 Calmette$).tw. (7331)
23. tubercul$ vaccin$.tw. (890)
24. Calmette Vaccin$.tw. (17)
25. or/20-24 (28239)
26. 19 and 25 (18248)
27. exp animals/ not humans/ (4527358)
28. 26 not 27 (13646)
29. limit 28 to yr="2010-current" (2497)

Table 1: Summary of cohort studies

| Location/ participants | Event at recruitment | Recruitment start, recruitment end, follow-up end | Estimated average follow-up period (years) | Age range or average age (years) | Case definition | Protective efficacy (confidence interval) | Estimate type |
|---|---|---|---|---|---|---|---|
| Edinburgh | Contact, vaccination | 1977, 1981, 1983 | 0.5 | All ages | TB | 0.51 (0.29, 0.88) | Crude RR |
| Seoul | Contact | 1984, 1986, 1983 | 0.25 | Under 5s | Scoring-system | 0.26 (0.18, 0.38) | Adjusted RR |
| Bangui | Contact | 1989, 1990, 1991 | 0.5 | Under 7s | Scoring-system | 0.29 (0.19, 0.44) | Crude RR |
| Edinburgh | Contact, vaccination | 1982, 1991, 1992 | 0.5 | All ages | TB | 0.45 (0.24, 0.84) | Crude RR |
| United Kingdom | Contact, vaccination | 1973, 1974, 1976 | 2 | All ages | TB | 0.31 (0.18, 0.54) | Crude RR |
| Oslo nurses | Nursing school entry, vaccination | 1924, 1936, 1946 | 3 | Adults | TB | 0.20 (0.14, 0.28) | Crude RR |
| Oslo | Vaccination | 1924, not stated, 1946 | Unclear | Not stated | TB | 0.20 (0.10, 0.40) | Crude RR |
| Richmond (USA) | Vaccination | Not stated | 7 | Under 6 months | TB | 0.01 (0, 0.05) | Crude RR |
| Rzeszow (Poland) | Population study | 1965, 1965, 1977 | 12 | Not stated | TB (unclear definition) | 0.22 (0.11, 0.47) | Crude RR |
| Trysil (Norway) | Vaccination | 1927, 1929, 1929 | 2 | Not stated | TB | 0.16 (0.03, 0.79)* | Crude RR |
| New York | Contact (family member), vaccination | 1926, 1932, 1932 | 6 | Zero to six | Not stated | 0.10 (0.01, 1.61) | Not stated |
| Bejaia (Algeria) schoolchildren | Vaccination | 1950, 1950, 1954 | 3 | 5 to 15 | CXR change | 0.55 (0.32, 0.93) | Crude RR |
| Northern France schoolchildren | Vaccination | 1948, 1951, 1971 | 20 | School children | CXR change | 0.27 (0.22, 0.33) | Crude RR |

| Location/ participants | Event at recruitment | Recruitment start, recruitment end, follow-up end | Estimated average follow-up period (years) | Age range or average age (years) | Case definition | Protective efficacy (confidence interval) | Estimate type |
|---|---|---|---|---|---|---|---|
| Karonga (Malawi) | Vaccination | 1979, 1984, 1984 | 5 | All ages | TB | 0.46 (0.32, 0.67) | Crude RR |
| Aker (Norway) | Vaccination | 1947, 1949, 1959 | 2.5 | All ages | CXR change | 0.54 (0.28, 1.06) | Crude RR |
| Virginia | Vaccination | 1948, 1948, 1968 | 10 | Infants | TB | 0.03 (0.00, 0.52) | Crude RR |
| Boston nurses | Nursing school entry, vaccination | 1947, 1951, 1951 | 4 | Adults | TB | 5.23 (0, >100) | Crude RR |
| Bornholm | Vaccination | 1936, 1946, 1946 | 9 | All ages | TB | 0.17 (0.05, 0.55) | Crude RR |
| Salvador (Brazil) | Entry to RCT (no revaccination) | 1996, 1998, 2007 | 9 | Neonates and children | Not stated | 0.6 (0.46, 0.78) | Crude RR |
| Manaus (Brazil) | None Entry to RCT (no revaccination) | 1998, 1998, 2007 | 9 | Neonates and children | Not stated | 0.64 (0.46, 0.89) | Crude RR |
| Salvador (Brazil) | Entry to RCT (no revaccination) | 1996, 1998, 2007 | 9 | Children and adolescents | Not stated | 0.66 (0.47, 0.93) | Crude RR |
| Manaus (Brazil) | Entry to RCT (no revaccination) | 1998, 1998, 2007 | 9 | Children and adolescents | Not stated | 0.89 (0.58, 1.35) | Crude RR |
| Lyon university students | University entry, vaccination | 1956, not stated, 1963 | Not stated | Young adults | TB | 0.17 (0.09, 0.31) | Crude RR |
| Norway | None | 1956, 1973, 1973 | 17 | Average 13 | TB | 0.18 (0.15, 0.23) | Crude RR |
| Dublin student nurses | Vaccination | 1933, 1949, 1954 | 5 | Average 18 | TB | 0.01 (0.00, 0.14) | Crude RR |
| Netherlands student nurses | Nursing school entry, vaccination | 1939, 1950, 1950 | Not stated | Young adults | TB | 0.15 (0.05, 0.50) | Crude RR |
| Districts of Hesse (Germany) | None | 1947, 1949, not stated | Not stated | Children | TB | 0.19 (0.14, 0.25) | Crude RR |
| Hamburg | None | 1950, 1971, not stated | Not stated | Under 15 | TB | 0.05 (0.03, 0.08) | Crude RR |
| Strasbourg university students | University entry | 1947, 1950, 1950 | Not stated | Young adults | CXR change and clinical | 0.07 (0.01, 0.65) | Crude RR |

| Location/ participants | Event at recruitment | Recruitment start, recruitment end, follow-up end | Estimated average follow-up period (years) | Age range or average age (years) | Case definition | Protective efficacy (confidence interval) | Estimate type |
|---|---|---|---|---|---|---|---|
| Philadelphia | Nursing school entry, vaccination | 1950, 1955, 1957 | 2 | Adults | Pulmonary TB | 0.27 (0.01, 5.37) | Crude RR |
| California medical students | Medical school entry | 1974, 1975, 1975 | 29 | Adults | TB | 0.55 (0.27, 1.15) | Crude RR |
| Jordan | Birth | 1980, 1986, 1994 | 11 | 0 | TB | 0.15 (0.1, 0.22) | Crude RR |
| Chicago medical students | Medical school entry, vaccination | 1982, 1982, 1982 | <1 | Not stated (adults) | TB | 0.6 (0.20, 1.86) | Crude RR |
| Uruguay | Hospital attendance | 1943, 1952, 1952 | Not stated | Zero to three | TB | 0.21 (0.13, 0.32) | Crude RR |
| UK medical students | Medical school entry, vaccination | 1950, 1953, 1953 | 3 | Young adults | TB | 0.03 (0, 0.11) | Crude RR |
| Germany siblings | Child contacts of TB cases | 1949, 1954, Not stated | Not stated | Children | TB | 0.002 (<0.001, 0.03) | Crude RR |
| Edinburgh school children | Vaccination | 1984, 1993, 1994 | 3.5 | Average age 13 | TB | 6.08 (0, >100) | Crude RR |
| Dusseldorf | Birth, vaccination | 1954, 1961, 1967 | 9 | 0 | TB | 0.09 (0.08, 0.11) | Crude RR |
| Ancona (Italy) | Contact (most participants) | 1938, 1940, 1955 | 16 | Not stated | Not stated | 0.37 (0.22, 0.61) | Crude RR |
| Morocco | Vaccination | 1950, 1951, not stated | 2 | One to 20 | CXR change with clinical features | 0.28 (0.18, 0.45) | Crude RR |
| Sweden conscripts | Vaccination | 1941, 1944, 1944 | 3 | Adults | Pulmonary TB | 0.37 (0.31, 0.45) | Crude RR |
| Norway deportees | Deportation to concentration camp | 1943, 1943, 1945 | 2 | Average age 24 | TB | 0.38 (0.13, 1.06) | Crude RR |
| Kazakhstan | Birth, vaccination | 2002, 2006, 2008 | 3 | 0 | Notified TB with radiological evidence | 0.54 (0.07, 0.62) | Crude RR |
| South Delhi | Contact | 2007, 2009, 2011 | 2 | Mean 27 | TB | 0.55 (0.31, 0.95) | Crude RR |

| Location/ participants | Event at recruitment | Recruitment start, recruitment end, follow-up end | Estimated average follow-up period (years) | Age range or average age (years) | Case definition | Protective efficacy (confidence interval) | Estimate type |
|---|---|---|---|---|---|---|---|
| Kampala | Household contact | 2002, 2012, 2014 | 2 | Median 13 | Secondary TB (ATS criteria) | 0.78 (0.58, 1.06) | Crude RR |
| British Columbia | Contact | 1990, 2000, 2002 | 12 | Median 35 | Author-defined TB | 0.74 (0.51, 1.03) | Crude RR |
| Worcester (South Africa) | None | 2005, 2007, 2009 | 2 | 12 to 18 | Bacteriologically-confirmed TB | 0.65 (0.36, 1.16) | Crude HR |
| Tasiilaq (Greenland) | Birth, vaccination | 1982, 2006, 2012 | 16 | 0 | WHO case definition | 0.35 (0.23, 0.52) | Crude RR |
| Siaya County (Kenya) | None | 2008, 2009, 2011 | 1.2 | 14 | TB | 1.13 (0.27, 4.80) | Crude RR |
| India | Suspected TB | 2010, 2012, 2012 | 1.5 | 2.6 | TB or suspicious radiology | 0.83 (0.25, 2.85) | Crude RR |
| Lima | Household contact | 2009, 2012, 2012 | 1 | All ages | TB | 0.77 (0.54, 1.09) | Crude RR |
| Norway | Vaccination | 1962, 1975, 2011 | 40 | 12 to 50 years | Notified TB | 0.38 (0.30, 0.49) | Crude RR |
| Uganda | Contact | Not stated | 2 | All ages | Culture-positive TB | 0.63 (0.39, 1.01) | Crude RR |
| UK | Contact | 2008, 2008, 2008 | $<1$ | 3 | TB | 0.32 (0.07, 1.52) | Crude RR |
| Amsterdam | Contact | 2002, 2011, 2011 | Not stated | Average 4.5 | Incident TB | 1.69 (1.16, 2.46) | Crude RR |
| Japan | None | 2005, 2008, 2008 | Not stated | Infants (0-3 years) | Incident TB | 0.29 (0.14, 0.61) | Crude RR |

* Indicates study with significant difference between our interpretation and that of Abubakar et al.

[37] A K Mehrotra, S B Dabral, D Nandan, B B Maheshwari, V Shrotriya, B Prakash, and S C Gupta. Assessment of efficacy of BCG vaccination among slum children of Agra City. *Indian journal of public health*, 32(3):164–5.

[38] K P Mehta, S M Merchant, and U Korde. Environmental influence on immunity due to BCG vaccination. *Indian pediatrics*, 13(7):525–32, jul 1976.

[39] I. S. Neiman and E. Loewinsohn. Inhibition of Primary Tuberculosis by BCG: A Study in Children, Based on 13,470 Chest Roentgenograms. *American Review of Tuberculosis*, 56(1):27–35, 1947.

[40] Elisa Nemes, Hennie Geldenhuys, Virginie Rozot, Kathryn T. Rutkowski, Frances Ratangee, Nicole Bilek, Simbarashe Mabwe, Lebohang Makhethe, Mzwandile Erasmus, Asma Toefy, Humphrey Mulenga, Willem A. Hanekom, Steven G. Self, Linda-Gail Bekker, Robert Ryall, Sanjay Gurunathan, Carlos A. DiazGranados, Peter Andersen, Ingrid Kromann, Thomas Evans, Ruth D. Ellis, Bernard Landry, David A. Hokey, Robert Hopkins, Ann M. Ginsberg, Thomas J. Scriba, and Mark Hatherill. Prevention of M. tuberculosis Infection with H4:IC31 Vaccine or BCG Revaccination. *New England Journal of Medicine*, 379(2):138–149, jul 2018.

[59] Tuberculosis Prevention Trial. Trial of BCG vaccines in south India for tuberculosis prevention: First report. Technical Report 5, 1979.

[60] Tuberculosis Vaccines and Clinical Trials Committee. BCG and vole bacillus vaccines in the prevention of tuberculosis in adolescents first (progress) report to the Medical Research Council by their tuberculosis vaccines clinical trials committee. Technical report, 1956.

[61] HM. M Vandiviere, M. Dworski, IG. G Melvin, KA. A Watson, and J. Begley. Efficacy of Bacillus Calmette-Guerin and isoniazid-resistant Bacillus Calmette-Guerin with and without isoniazid chemoprophylaxis from day of vaccination. *American Review of Respiratory Disease*, 108(2):301–13, aug 1973.

[62] World Health Organization. Global immunization profile, 2019.

[63] Alice Zwerling, Marcel A. Behr, Aman Verma, Timothy F. Brewer, Dick Menzies, and Madhukar Pai. The BCG World Atlas: A Database of Global BCG Vaccination Policies and Practices. *PLoS Medicine*, 8(3):e1001012, mar 2011.
